# Effects of Cannabidiol on Social Relating, Anxiety, and Parental Stress in Autistic Children: A Randomized Controlled Crossover Trial

**DOI:** 10.1101/2024.06.19.24309024

**Authors:** Nina-Francesca Parrella, Aron T. Hill, Peter G. Enticott, Tanita Botha, Sarah Catchlove, Luke Downey, Talitha C. Ford

## Abstract

Cannabidiol (CBD), a non-intoxicating compound derived from the cannabis plant, has garnered increasing attention as a potential pharmacological therapeutic for autism. We conducted a randomized, double-blind, placebo-controlled, crossover trial to understand whether oral CBD oil containing terpenes can improve outcomes in autistic children. Twenty-nine children (18 male), aged 5 to 12 years (M=9.62 years, SD=2.05), diagnosed with autism spectrum disorder, completed the study. Participants received weight-based dosing of CBD oil (10 mg/kg/day) or matched placebo oil over two 12-week intervention periods (crossover), separated by an 8-week washout period. Outcome measures included the Social Responsiveness Scale-2 (SRS-2; primary outcome), PROMIS Social Relating, Anxiety, and Sleep, Developmental Behaviour Checklist-2 (DBC-2), Vineland-3, and Autism Parenting Stress Index (APSI; secondary outcomes). There was no significant effect observed for the primary outcome measure (SRS-2) for CBD oil relative to placebo oil after 12 weeks (β=-11.15, SE=7.19, *p*=.125). Significant improvements were observed in secondary measures of social functioning, including DBC-2 Social Relating (β=-2.35, SE=0.92, *p*_(adj)_=.024), as well as reduced anxiety on the DBC-2 subscale (β=-3.20, SE=0.94, *p*_(adj)_=.002), and lower parental stress (APSI; β=-4.63, SE=2.26, *p*_(adj)_=.044). No differences were detected on Vineland-3 adaptive functioning (ABC: β=2.06, SE=2.67, *p*_(adj)_=1.000), and domain scores were not significant. Safety and tolerability data indicated that two children experienced gastrointestinal discomfort while taking CBD. Findings from this pilot trial suggest that while CBD combined with terpenes did not improve the primary outcome of social responsiveness, it may hold potential in addressing certain autism-related difficulties, particularly anxiety and social relating. Further research with larger sample sizes is needed to fully evaluate the efficacy and safety of CBD for autistic children.

**Lay Summary:** This pilot, exploratory study examined whether taking CBD oil could improve social outcomes for autistic children. While no significant changes were seen in overall social responsiveness, CBD led to improvements in specific social behaviours, reduced anxiety, and lowered parental stress. The treatment was generally well tolerated, though a few children experienced mild side effects. These findings suggest potential benefits of CBD for some autism-related symptoms, but larger studies are needed to confirm effectiveness.

## Introduction

Autism is characterized by core social communication/interaction difficulties (American Psychiatric Association, 2022), with frequently co-occurring anxiety and elevated parental stress (Buchwald et al., 2025; Thiele-Swift & Dorstyn, 2024). Although GABAergic approaches show early promise (Veenstra-VanderWeele et al., 2017), no pharmacological treatments are currently approved for social functioning in autism. In Australia, risperidone is the only approved medicine for behavioral disorders in autistic children/adolescents (Janssen-Cilag Pty Ltd, 1993), while other antipsychotics, antidepressants/anxiolytics, and stimulants are typically used off-label but show limited efficacy and can cause adverse side effects (Baldes et al., 2024; Manter et al., 2025). Current support largely relies on behavioral approaches, which often carry a substantial time cost and burden. Cannabidiol (CBD) is a non-intoxicating, non-addictive compound derived from the *Cannabis sativa (C. sativa)* L. plant with emerging therapeutic potential for various somatic, psychiatric, and neurodevelopmental conditions, including autism (Aran et al., 2021; Larsen & Shahinas, 2020; VanDolah et al., 2019). Given the unmet need for safe and effective therapeutic options for autistic individuals, the present study focuses on CBD intervention with core social difficulties and associated domains (anxiety, parental stress) as clinically meaningful targets.

The endocannabinoid system (ECS) is a neuro-modulatory network involved in regulating social behavior, anxiety, and sleep, with preliminary evidence of relevance to autism (Karhson et al., 2016; Wei et al., 2016). Preclinical studies indicate that changes to ECS signaling can produce autism-like social and repetitive phenotypes, and restoring signaling can normalize social behavior in animal models (Wei et al., 2016; Zamberletti et al., 2017). Emerging data also suggests ECS dysregulation in patients with autism, including alterations in endocannabinoid-linked pathways related to social function (Karhson et al., 2016, 2018). These findings support the ECS as a plausible therapeutic target in autism, and have motivated early clinical exploration of pharmacological approaches that modulate the ECS to support social communication difficulties (Karhson et al., 2016; Wei et al., 2016).

CBD has multiple pharmacological properties, which include anti-inflammatory (Weiss et al., 2008), anxiolytic (Bergamaschi et al., 2011), neuroprotective and pro-cognitive effects (Osborne et al., 2017). There has been a notable increase in the use of CBD for psychiatry-related conditions in Australia (Cairns et al., 2023), alongside a surge in research exploring therapeutic benefits (Abi-Jaoude et al., 2022; Cooper et al., 2016). Mechanistically, CBD likely engages endocannabinoid and related neuro-modulatory systems, though precise therapeutic pathways remain unclear (Gunasekera et al., 2020). There is growing scientific interest in CBD as a potential therapeutic for neurodevelopmental disorders due to its non-intoxicating nature and robust safety profile (Chin et al., 2020; Lattanzi et al., 2021), although existing clinical trials show variable rigor regarding safety and efficacy (Parrella et al., 2023).

Emerging research suggests potential benefits of CBD for autism-related difficulties, and there is evidence for reductions in disruptive behaviours and anxiety following multi-week administration (Aran et al., 2021; Efron et al., 2021; Silva Junior et al., 2024). There is, however, significant variability in CBD study findings for autism and an absence of standardized approaches and dosage reporting across RCTs in this field (Parrella et al., 2023). For example, study designs include open-label assessments, as well as placebo-controlled parallel and crossover designs, with sample sizes ranging from 8 to 150 participants (Aran et al., 2021; Efron et al., 2021; Heussler et al., 2022). Three trials specifically included predominantly children (Aran et al., 2021; Efron et al., 2021; Silva Junior et al., 2024). Dosage regimens and administration methods/routes also vary across trials, as do primary and secondary outcome measures (Heussler et al., 2022; Silva Junior et al., 2024). Although the literature includes studies in autistic adults and acute/single-dose paradigms, these have limited relevance to pediatric clinical practice and to the sustained social-communication goals of families and clinicians (Pretzsch, Freyberg, et al., 2019; Pretzsch, Voinescu, et al., 2019). There are 12 ongoing RCTs examining the long-term effects of CBD in neurodevelopmental disorders, which vary in trial duration (from 6 to 34 weeks), but do adopt similar methodologies, facilitating future comparison (Parrella et al., 2023). The existing studies highlight the need for further research to confirm the efficacy and safety of CBD in supporting social communication, a core diagnostic domain of autism.

Given the lack of studies focusing on social outcomes, we conducted a Phase II randomized, double-blind, placebo-controlled crossover trial to investigate whether oral CBD oil improves social relating in autistic children. Secondary aims were to evaluate CBD effects on anxiety, parental stress, and other social and adaptive behavior measures over 12 weeks. Our primary hypothesis was that 12-week administration of CBD oil would positively impact social outcomes for autistic children, as measured by the Social Responsiveness Scale – 2^nd^ Edition (SRS-2). Specifically, we hypothesized that participants would exhibit a significant reduction in SRS-2 scores from baseline to 12 weeks. Safety, tolerability, and acceptability data were also collected. To our knowledge, this is the first double-blind, placebo-controlled crossover RCT to evaluate a ‘broad-spectrum’ CBD plus terpenes oil extract (with negligible THC) in Australian autistic children.

## Methods

### Ethics, Consent, and Registration

This trial was prospectively registered under the Australian New Zealand Clinical Trials Registry (<u>ACTRN12622000437763</u>) and received ethics approval from the Deakin University Human Research Ethics Committee (DUHREC 2020-071) prior to enrolment of the first participant. Written informed consent was obtained from the parents/guardians of all participants prior to participation in the study, and assent was obtained from participants when appropriate. The only deviation from the protocol was that adherence to the daily Treatment Log was lower than anticipated for some participants.

### Trial Design

This randomized, double-blind, placebo-controlled crossover trial was conducted at Deakin University’s School of Psychology in Melbourne, Australia. For each participant, the trial spanned a total of 32 weeks, divided into two 12-week intervention periods, with an 8-week washout period in between. Eligible participants were children (5-12 years old) with an autism spectrum disorder (ASD) diagnosis per DSM-5 criteria. Evidence of diagnosis was obtained from the participant’s clinician. The trial aimed to investigate the effects of weight-based dosing of a medicinal cannabis oil product (Cannabidiol: Medigrowth CBD100) on social relating in autistic children. There were no changes made to eligibility criteria once the trial had commenced. No scientific or methodological amendments were made after enrolment began. One administrative amendment (ethics approval 14 Aug 2023) authorised the use of Calendly to schedule visits during Period 2 of the trial. This had no impact on eligibility criteria, interventions, outcomes, or the statistical plan.

### Participants

Thirty-four children were enrolled in the study. Five participants withdrew their participation during the first intervention period, leaving a total of 29 participants completing the trial. ASD diagnoses (per DSM-5) were established prior to enrolment by external specialist clinicians and verified by the study team at screening via review of clinical reports and/or multidisciplinary letters. No study-administered diagnostic re-verification (e.g., ADOS-2/CARS-2) was undertaken due to feasibility and participant-burden considerations in this pilot.

Formal IQ testing was not part of the study protocol; therefore, IQ data are not reported. Inclusion and exclusion criteria for study participation is in Table 1, participant demographics in Table 2, and participant medications in Table 3.

**Table 1.**
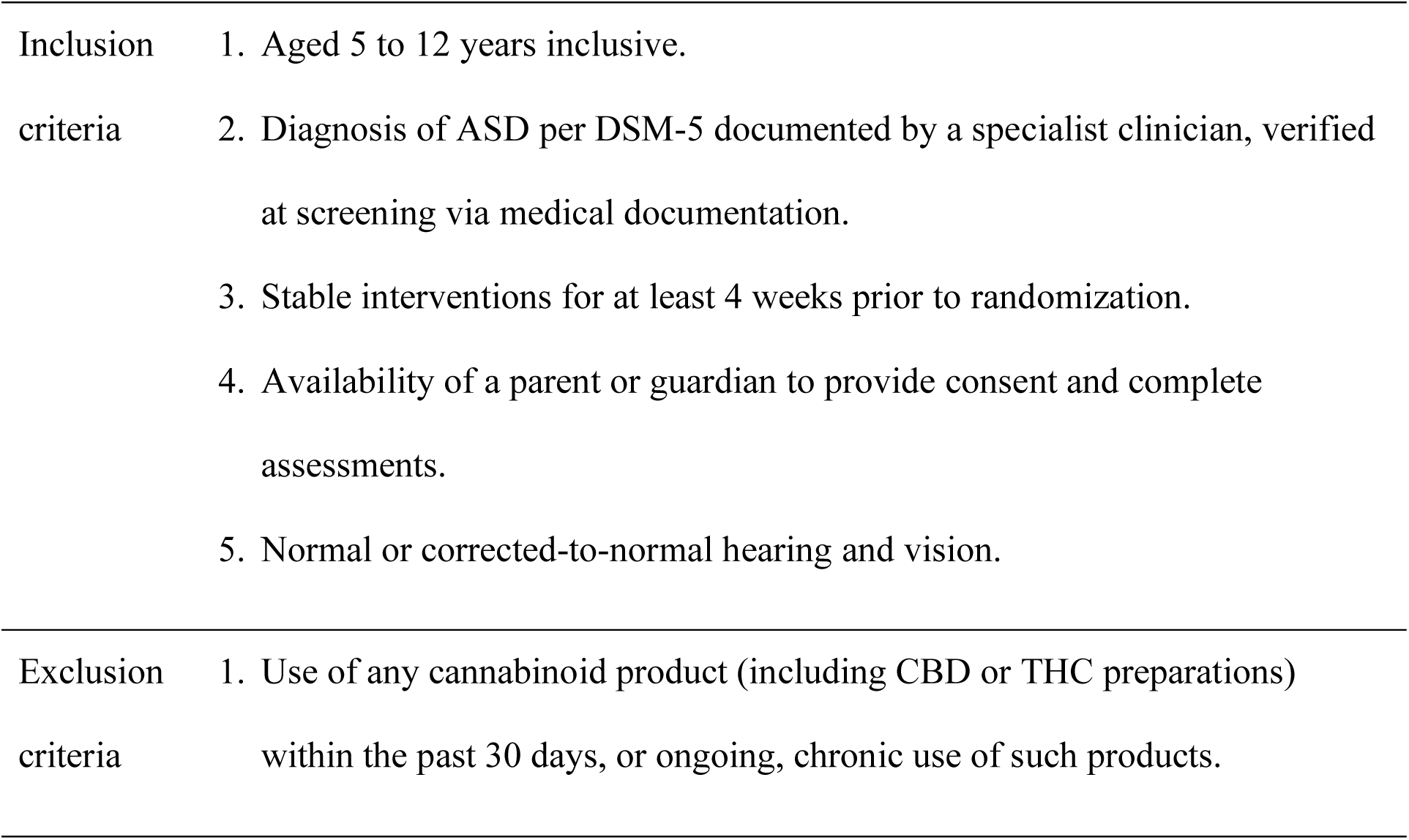
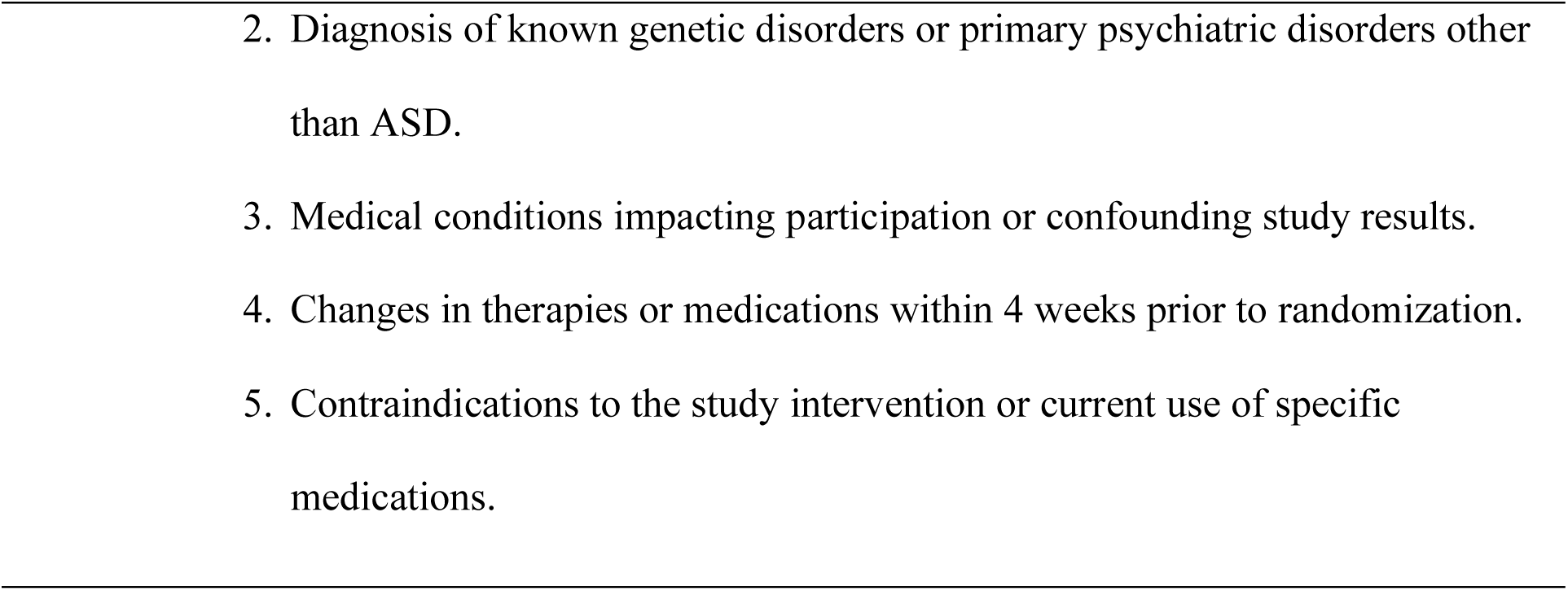
Inclusion and exclusion criteria.

**Table 2.**
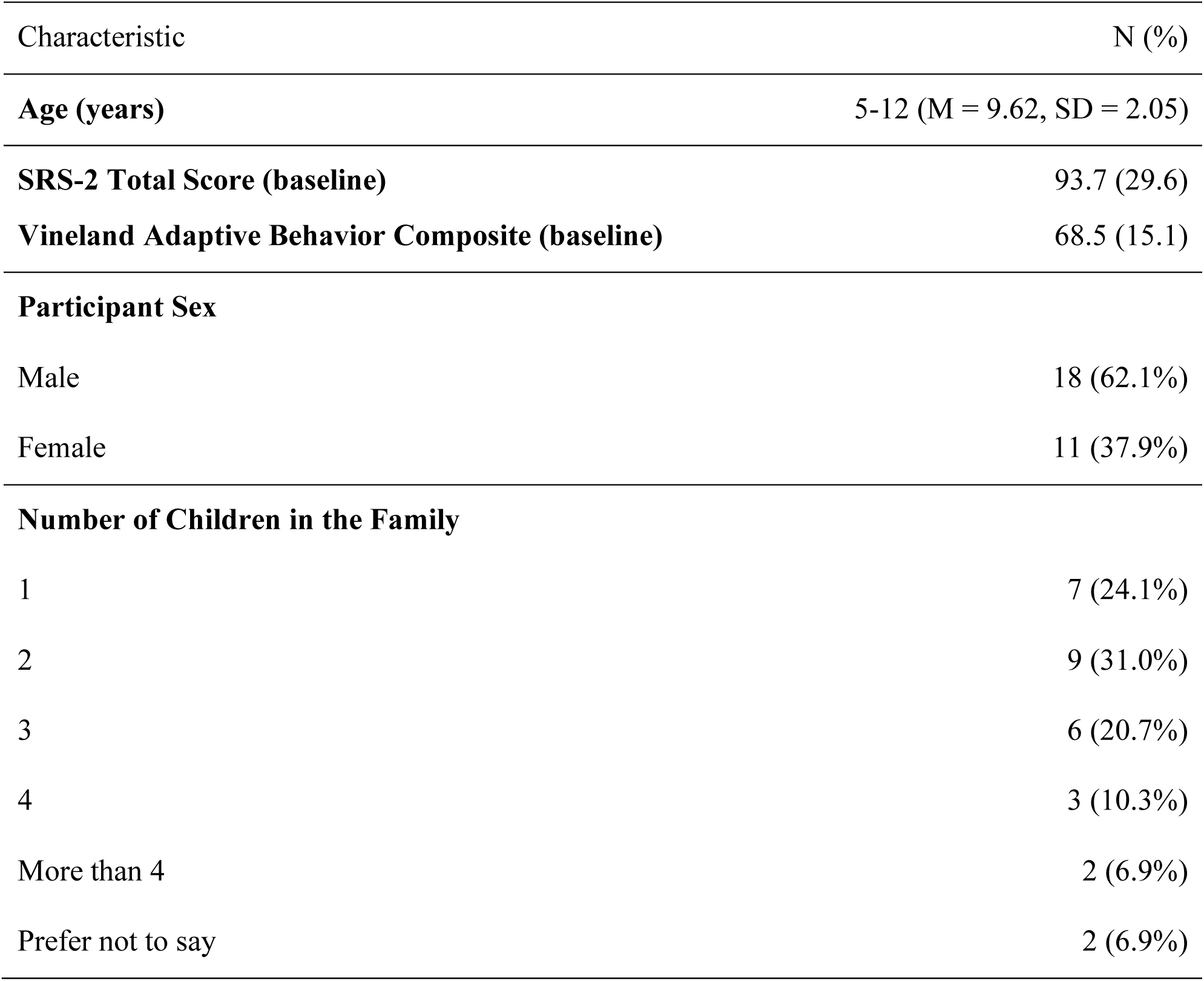
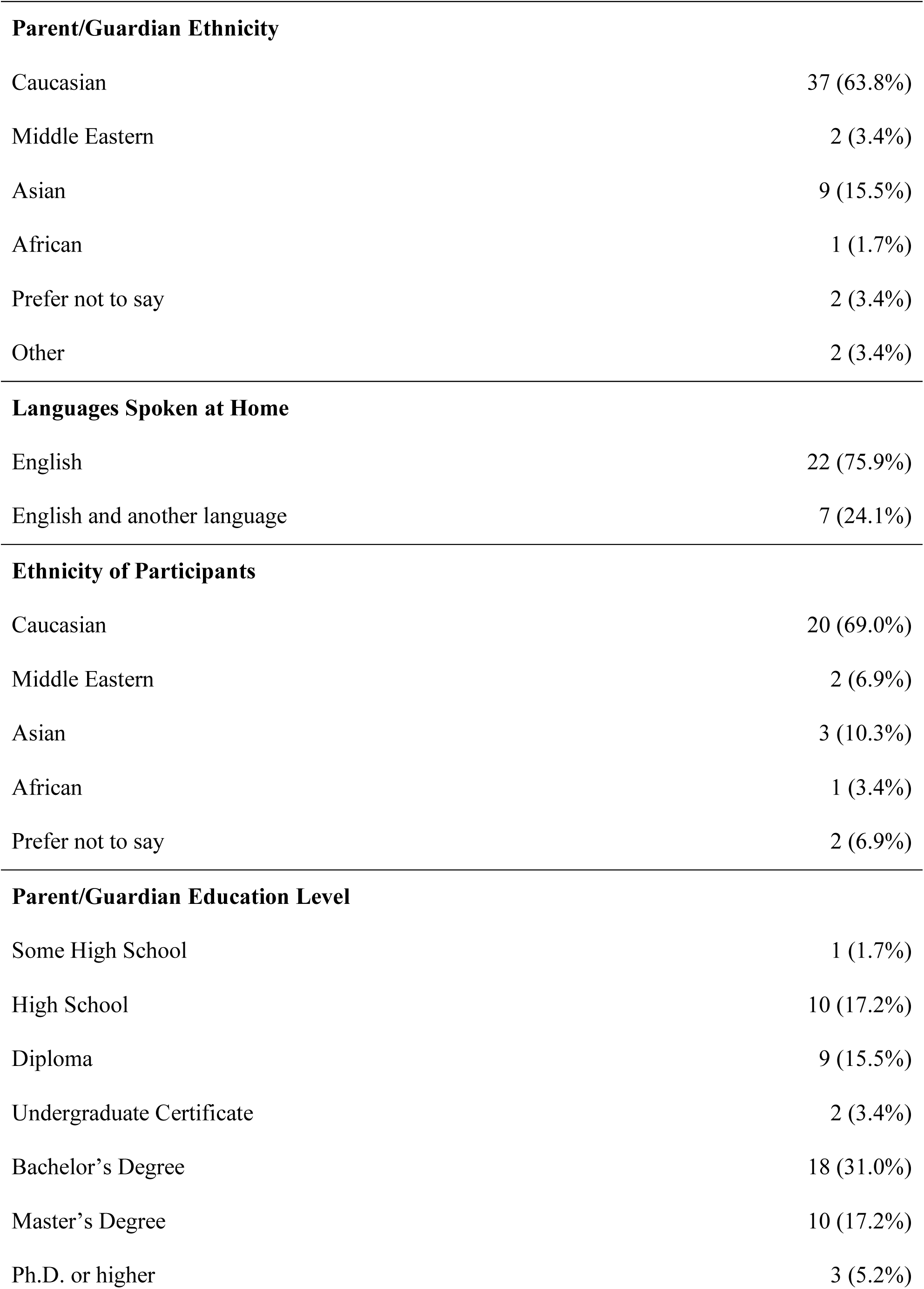
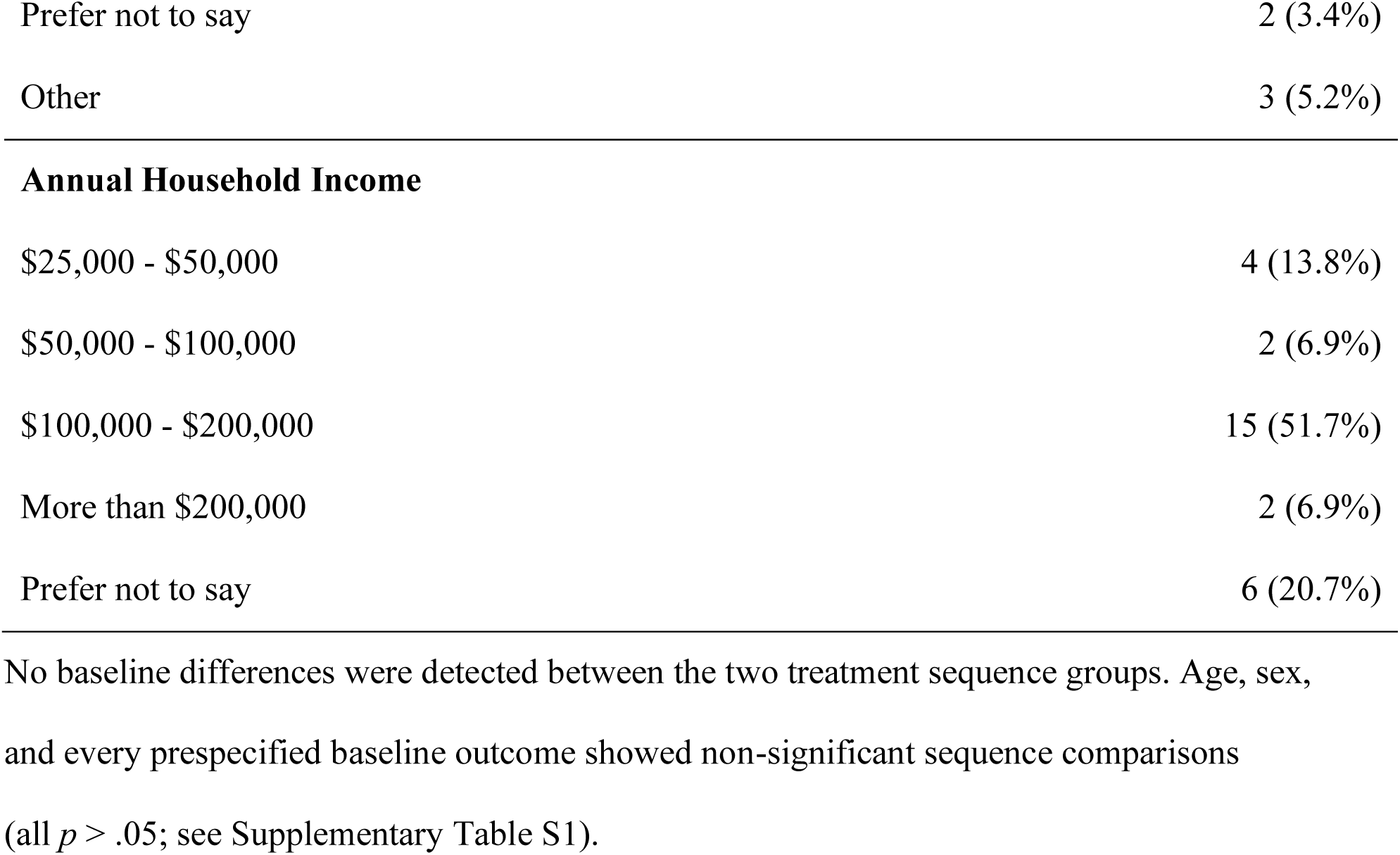
Demographics of participants and their families (N = 29)

| Characteristic | N (%) |
| --- | --- |
| **Age (years)** | 5-12 (M = 9.62, SD = 2.05) |
| **SRS-2 Total Score (baseline)** | 93.7 (29.6) |
| **Vineland Adaptive Behavior Composite (baseline)** | 68.5 (15.1) |
| **Participant Sex** |  |
| Male | 18 (62.1%) |
| Female | 11 (37.9%) |
| **Number of Children in the Family** |  |
| 1 | 7 (24.1%) |
| 2 | 9 (31.0%) |
| 3 | 6 (20.7%) |
| 4 | 3 (10.3%) |
| More than 4 | 2 (6.9%) |
| Prefer not to say | 2 (6.9%) |
| **Parent/Guardian Ethnicity** |  |
| Caucasian | 37 (63.8%) |
| Middle Eastern | 2 (3.4%) |
| Asian | 9 (15.5%) |
| African | 1 (1.7%) |
| Prefer not to say | 2 (3.4%) |
| Other | 2 (3.4%) |
| **Languages Spoken at Home** |  |
| English | 22 (75.9%) |
| English and another language | 7 (24.1%) |
| **Ethnicity of Participants** |  |
| Caucasian | 20 (69.0%) |
| Middle Eastern | 2 (6.9%) |
| Asian | 3 (10.3%) |
| African | 1 (3.4%) |
| Prefer not to say | 2 (6.9%) |
| **Parent/Guardian Education Level** |  |
| Some High School | 1 (1.7%) |
| High School | 10 (17.2%) |
| Diploma | 9 (15.5%) |
| Undergraduate Certificate | 2 (3.4%) |
| Bachelor's Degree | 18 (31.0%) |
| Master's Degree | 10 (17.2%) |
| Ph.D. or higher | 3 (5.2%) |
| Prefer not to say | 2 (3.4%) |
| Other | 3 (5.2%) |
| **Annual Household Income** |  |
| \$25,000 - \$50,000 | 4 (13.8%) |
| \$50,000 - \$100,000 | 2 (6.9%) |
| \$100,000 - \$200,000 | 15 (51.7%) |
| More than \$200,000 | 2 (6.9%) |
| Prefer not to say | 6 (20.7%) |

**Table 3.**
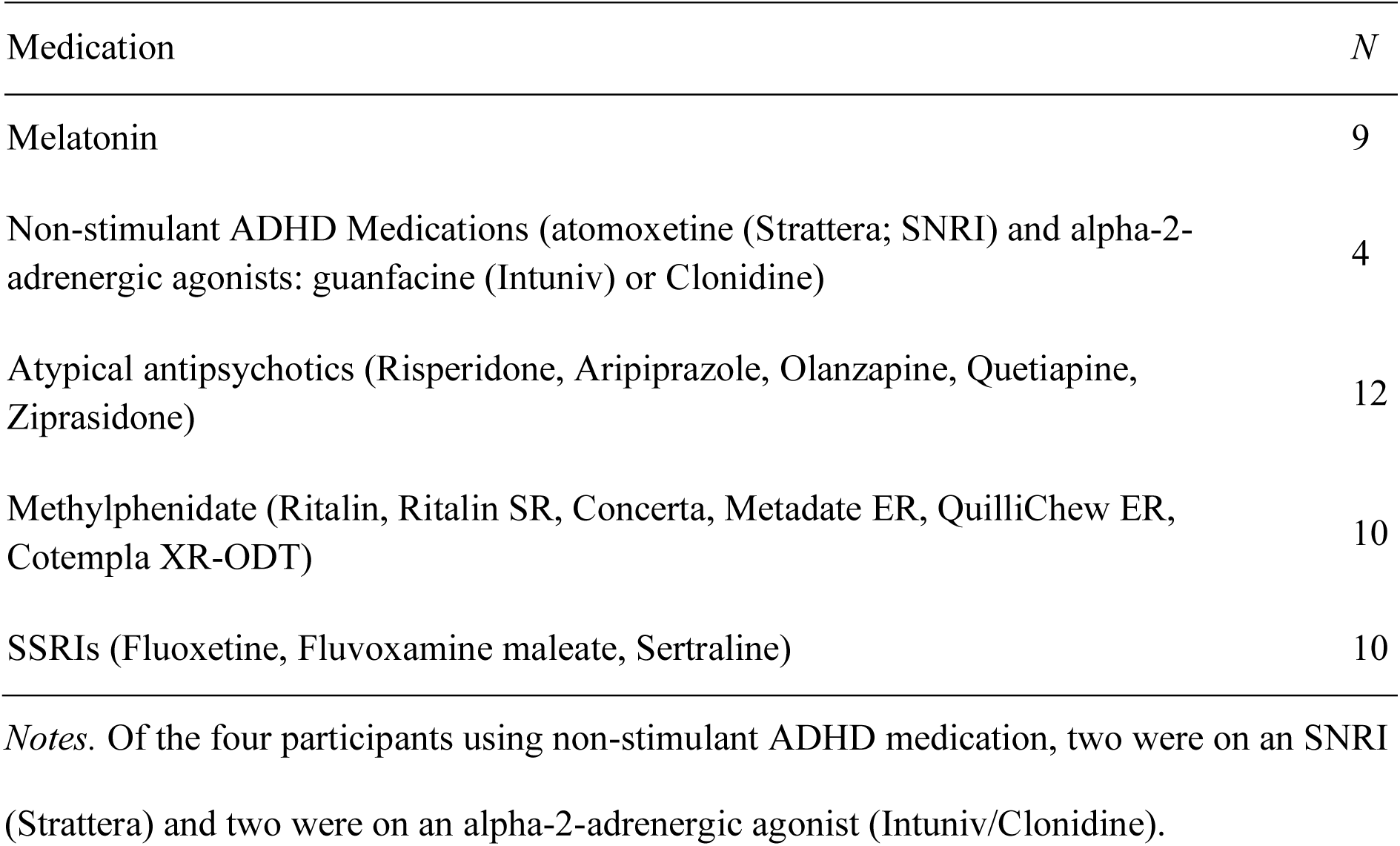
Existing participant medications maintained throughout trial.

### Intervention

Medigrowth CBD100 oil was distributed in a 30 mL bottle, with each milliliter of oil containing 100 mg of cannabidiol (CBD) and a negligible amount of Delta-9-trans-Tetrahydrocannabinol (THC) not exceeding 1 mg/mL. CBD100 is formulated with medium-chain triglyceride (MCT) excipients and naturally occurring terpenes including Nerolidol, α-Bisabolol, d-Limonene, β-Caryophyllene, and α-Humulene. The total cannabinoid content per bottle was 3000 mg. In addition, the placebo for the CBD100 medication consisted of MCT as an excipient, matched to CBD100 in taste to maintain blinding integrity throughout the trial. Taste and smell similarity between the CBD and placebo oils was established during product development and confirmed through informal testing with independent staff not involved in the trial. Although no formal blinding assessment was conducted, procedures were designed to minimize the potential for unblinding by participants and caregivers.

Participants were randomized 1:1 to the two sequences (CBD-Placebo vs Placebo-CBD) using a computer-generated block randomization scheme with a block size of 4. Given the 8-week washout and balanced allocation, carryover was considered unlikely a priori. Nevertheless, we formally evaluated potential carryover using the Sequence × Period interaction in a mixed-effects model (Table 6). Baseline equivalence by sequence is shown in Supplementary Table S1. The allocation sequence was generated by a senior researcher on the team and stored in a secure electronic file accessible only to the designated team member responsible for assignment. Both the active CBD oil and the placebo were prepared to be identical in appearance, taste, and packaging, ensuring that participants, caregivers, and outcome assessors remained blinded throughout the trial.

Medigrowth CBD100 was administered orally in weight-based doses calculated by the investigator. Parents/guardians received in-person training at T0 on dose administration (syringe demonstration, timing, and recording procedures). Full step-by-step instructions are provided in Supplementary Methods S1 (Parent Administration Instructions). In each intervention period, the starting dose was 5 mg/kg body weight per day. The dose was increased to 10 mg/kg/day by the second week, with parents or guardians trained to facilitate sublingual administration during the first visit. We adopted this schedule as it sits within the pediatric safety range established in epilepsy trials (Devinsky et al., 2018; Thiele et al., 2018) and aligns with the CBD doses used in other RCTs involving children with autism (Aran et al., 2021; Efron et al., 2021), allowing cross-study comparison. Placebo oil was administered following the same protocol. Intervention adherence was monitored through a daily Treatment Log. Interventions were given as an add-on to any ongoing stable medication (no change in four weeks prior to study entry).

### Measures

#### Primary Outcome

The primary outcome was the *Social Responsiveness Scale – Second Edition* (SRS-2; Constantino & Gruber, 2012), a 65-item parent-report questionnaire used to assess the severity of autism spectrum symptoms in naturalistic social contexts. It comprises five subscales:

● Social Awareness: the ability to pick up on social cues.
● Social Cognition: the ability to interpret social signals.
● Social Communication: expressive and receptive communication skills in social contexts.
● Social Motivation: the level of interest in engaging with others socially.
● Restricted Interests and Repetitive Behavior (RRBs): the presence and severity of repetitive or stereotyped behaviours. Total and subscale scores were used in analyses.

The primary outcome was the Social Responsiveness Scale-Second Edition (SRS-2) Total T-score. In addition to the SRS-2 Total T-score (primary outcome), we also examined the Social Communication Index (SCI) and individual subscales to explore whether specific domains of social functioning were differentially affected by CBD. The SCI is a composite domain score derived from the Social Awareness, Social Cognition, Social Communication, and Social Motivation subscales, and is reported alongside the individual subscales to provide a more detailed understanding of potential treatment effects.

#### Secondary Outcomes

● Vineland Adaptive Behavior Scales – Third Edition (Vineland-3; Sparrow et al., 2016): A semi-structured caregiver interview that assesses adaptive functioning in Communication, Daily Living Skills, and Socialization domains.
● Developmental Behavior Checklist – Second Edition (DBC-2; Einfeld & Tonge, 2002): A parent-report tool assessing behavioral and emotional disturbance in children with developmental disabilities, including specific subscales for social relating and anxiety.
● Autism Parenting Stress Index (APSI; Silva & Schalock, 2012): A parent-report questionnaire that measures stress levels specifically related to parenting a child with autism.
● PROMIS Early Childhood Parent-Report – Social Relationships (PROMIS Health Organization and Assessment Center, 2014c): Captures the quality of the child’s social interactions and connectedness, as observed by parents.
● PROMIS Early Childhood Parent-Report – Anxiety (PROMIS Health Organization and Assessment Center, 2014a): Evaluates parental observations of a child’s anxiety symptoms.
● PROMIS Early Childhood Parent-Report – Sleep Health (PROMIS Health Organization and Assessment Center, 2014b): Assesses sleep-related behaviors and difficulties in young children.
● Repetitive Behavior Scale – Revised (RBS-R; Bodfish et al., 2000): A caregiver-report measure assessing the breadth and severity of repetitive behaviours in children with autism.
● Pediatric Quality of Life Inventory (PedsQL; Varni et al., 1999): A parent-report measure assessing children’s health-related quality of life across physical, emotional, social, and school functioning.
● Behavior Rating Inventory of Executive Function – Second Edition (BRIEF-2; Gioia et al., 2015): Evaluates executive functioning in everyday settings, as reported by caregivers.
● Personal Wellbeing Index – School Children (PWI-3; Cummins & Lau, 2005): A self-report measure evaluating children’s subjective wellbeing across multiple life domains.

To reduce measurement redundancy and align outcomes with our domains of interest, we selected nine outcomes for secondary analysis: PROMIS Social Relationships, DBC-2 Social Relating, PROMIS Anxiety, DBC-2 Anxiety, Vineland-3 Adaptive Behavior Composite (ABC), Vineland-3 Communication, Vineland-3 Daily Living Skills, Vineland-3 Socialization, and APSI. All other collected subscales from these instruments are treated as exploratory and are summarized descriptively in the Supplementary Table S2. Informant-based measures were completed by the same primary caregiver at each timepoint. The study coordinator confirmed respondent identity at each visit, provided clear, verbal instructions, and reviewed forms for completeness.

### Sample Size

A power analysis was conducted using the software package, G*Power 3 (Faul et al., 2007). The alpha level used for this analysis was *p* < .05. A sample size of 34 was determined for the detection of a moderate-large effect size (f^2^ = .25, power = .8). The determined sample size is consistent with a study involving acute administration of CBD in autistic individuals (Pretzsch, Freyberg, et al., 2019c), reflecting our best estimate given the limited available trials for comparison.

### Recruitment

Recruitment took place between January and March 2023. During the first intervention period of the trial, an attrition rate of 14.7% reduced the sample from 34 to 29. One participant was lost to follow-up, and four withdrew their participation, discontinuing intervention. Reasons for withdrawal included family-related difficulties for three participants (two families) and gastrointestinal discomfort for one other. Three of the four participants who discontinued their involvement were allocated the active intervention in the first intervention period.

### Procedure

Participants underwent an initial telephone screening to confirm eligibility and were then enrolled in the trial, comprising two intervention periods separated by an 8-week washout period, see Figure 1. Each period involved administration of either Medigrowth CBD100 or placebo oil, with the order of intervention randomized. Four testing visits took place throughout the trial. The first visit, T0, served as the baseline assessment for intervention period one, followed by T1 at 12 weeks. T2 marked the baseline assessment for intervention period two, with T3 occurring at 12 weeks thereafter, concluding the study’s assessment timeline.

**Figure 1.**
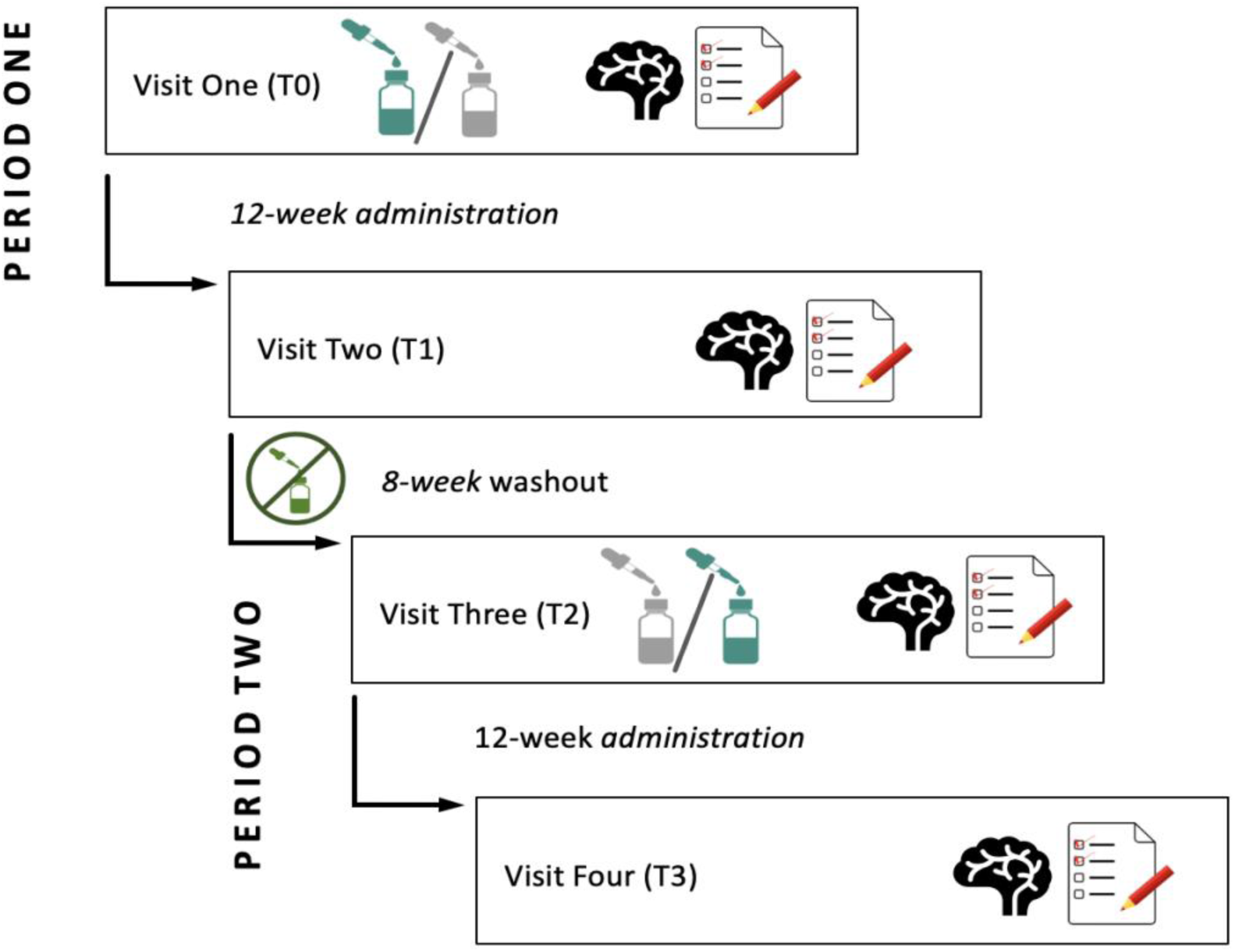
Study design illustrating two 12-week administration periods separated by an 8-week washout. Each period includes baseline and follow-up assessments with EEG recordings (brain icon) and behavioral (parental) questionnaires (paper icon).

Given the lack of pharmacokinetic data for CBD in children, an 8-week interval ensured adequate washout time for CBD100, which is at least four weeks (Thiele et al., 2018). The placebo condition served as the within-participants control measure to investigate CBD-specific outcomes in behaviour.

During the testing visits, parents or guardians of participants were required to complete behavioral questionnaires (Table 4). The SRS-2 and BRIEF 2 were administered in paper format, while all other measures were completed electronically via REDCap. The Vineland-3 and the DBC-2 were completed through their respective psychological assessment publisher platforms. Additionally, a post-study survey, administered via REDCap, collected economic information from the parents or guardians. The trial also involved a cohort of participants undergoing electroencephalography (EEG) recording to capture aperiodic neural activity. We conducted 5 minutes of eyes-open resting state EEG (64-channel), which will be reported separately.

**Table 4.**
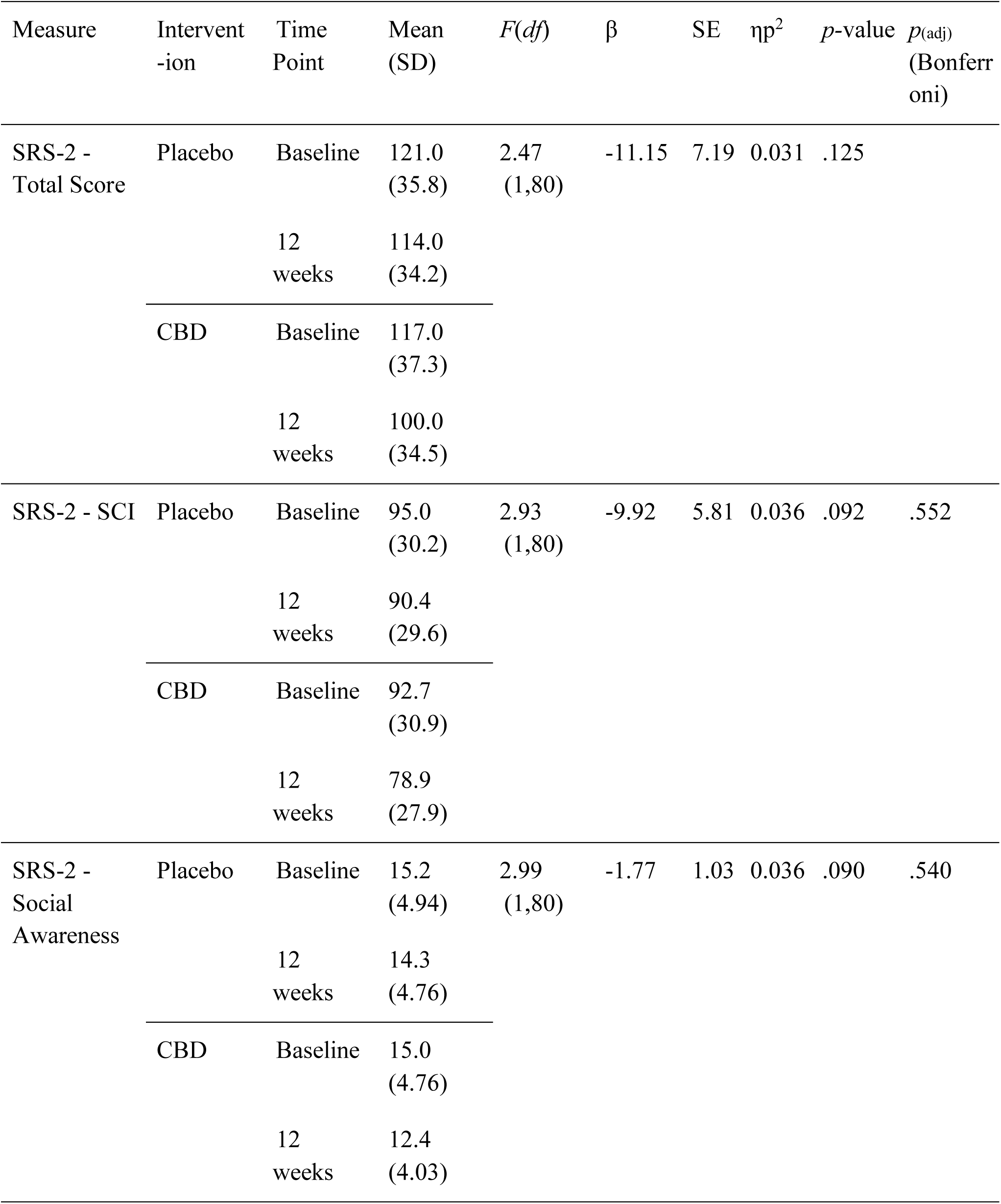
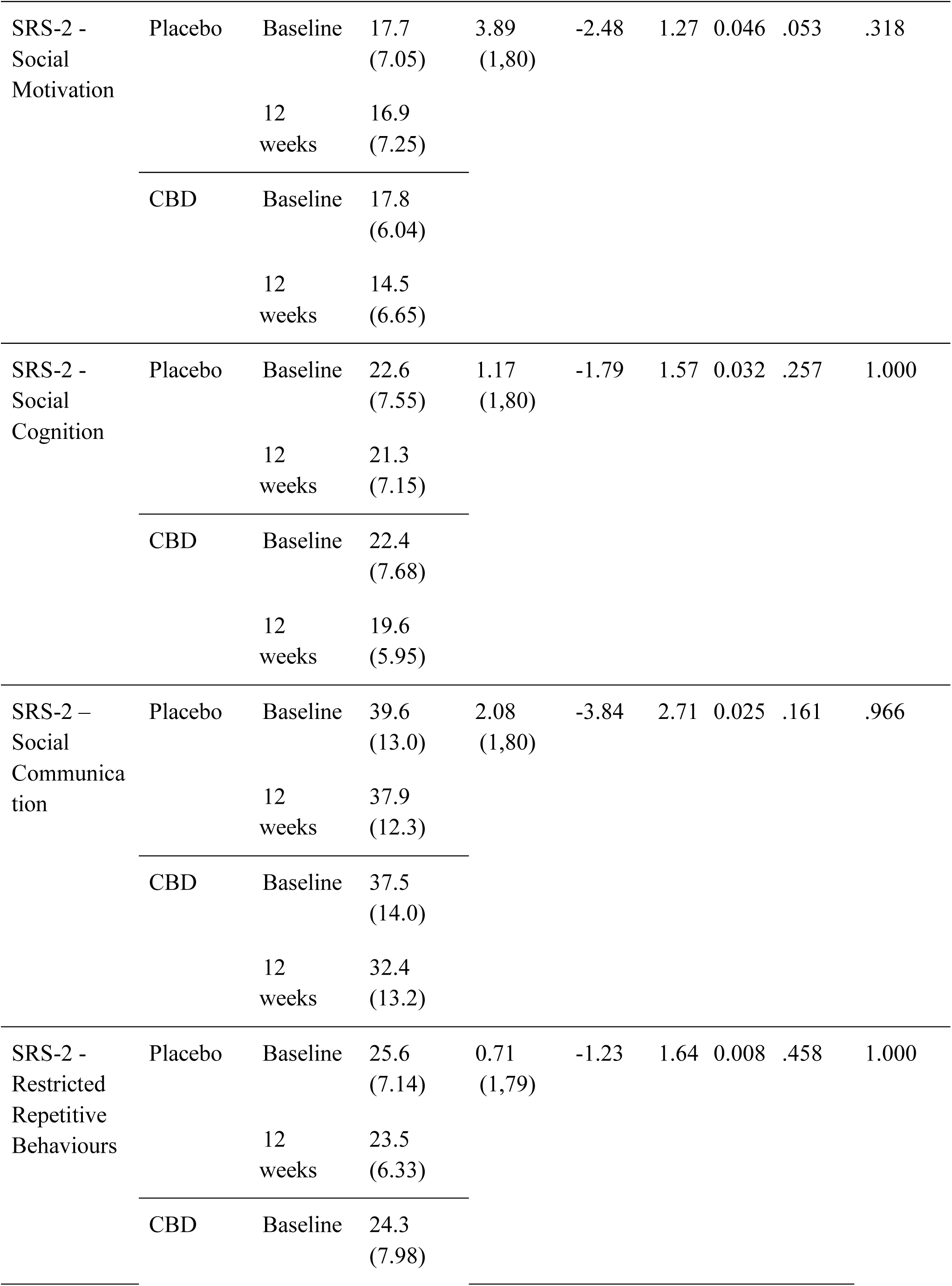
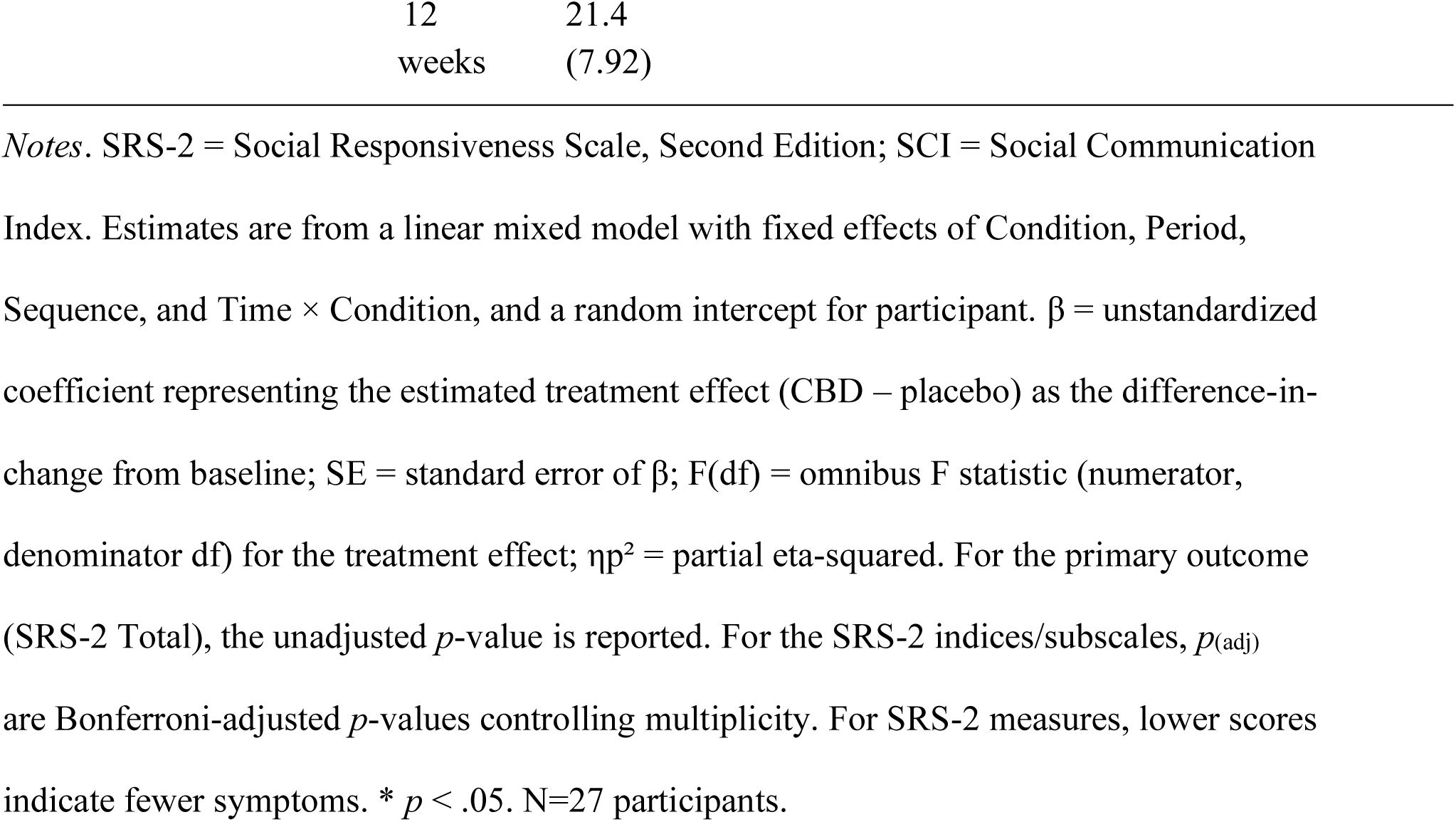
Summary of intervention effects on primary outcome measure (SRS-2 – Total Score), and composite and subscale scores from baseline to 12-weeks.

Parents or guardians of participants provided ASD DSM-5 diagnostic reports, completed behavioral questionnaires, and administered the study intervention daily. They also filled out a daily Treatment Log and assisted their children during testing visits. The online Treatment Log captured detailed information on administration, adherence, and child wellbeing. At T0 the researcher demonstrated administration of the calculated daily dose and advised splitting the total into two administrations (morning and evening) at consistent times. Parents recorded each dose in the daily Treatment Log (see Supplementary Methods S1). Specifically, the log recorded whether the oil was taken (and how much), the time(s) of administration, whether it was consumed with food (and what food), and any administration-related issues. It also included items assessing the child’s appetite and behavioral presentation, including a simplified emotion regulation tool using a “Feelings Thermometer” framework. Parents indicated which emotional “zones” their child experienced that day (e.g., Green = calm, Yellow = excited, Blue = withdrawn, Red = angry). Open-ended prompts allowed for reporting of any adverse events, concerns, or notable behavioral observations. A copy of the Treatment Log is included in the Supplementary Form S1.

### Statistical Analysis

Data analysis was conducted using mixed models within the Jamovi statistical software package (version 2.5.3). Mixed models were chosen due to their ability to account for within-subject correlation in crossover designs while accommodating fixed and random effects. For the primary analysis, a mixed-effects model was fitted to the outcome variable (SRS-2 Total, SCI), with Time (Baseline, 12 Weeks) and Intervention (CBD, placebo) as fixed effects. Participant was included as a random effect to account for within-subject correlation. The following parameters are reported: β (unstandardized coefficient), which represents the estimated treatment effect, i.e. the difference in mean change from baseline between the CBD and placebo periods; SE (standard error of β); *F*(*df*), the omnibus F-statistic for the treatment effect with its numerator and denominator degrees of freedom in parentheses; and ηp² (partial eta-squared), which expresses effect size as the proportion of outcome variance uniquely attributable to the Intervention factor after controlling for other terms in the model. Bonferroni was applied for multiple comparisons between primary outcome variables (*p*_(adj)_).

Multiplicity was also controlled using a Bonferroni adjustment for secondary outcomes, within prespecified outcome domains (Anxiety; Social Functioning; Caregiver Stress; Adaptive Functioning/Vineland-3). Adjusted *p*-values, *p*_(adj)_, are reported for secondary outcomes unless otherwise specified (α = .05). The estimated intervention effect along with 95% confidence intervals and associated *p-*values were reported. The assumption of normality was assessed visually through histograms and Q-Q plots and confirmed by the Shapiro-Wilk test. Two primary outcome observations showing implausible change between consecutive time points were excluded after blinded data checks. Both observations came from siblings within the same family, rated by the same primary caregiver, and the exclusion decision was made prior to unblinding. All analyses were conducted blind to the intervention condition, ensuring objectivity and minimizing potential bias in the interpretation of results. No missing data occurred for any outcome. Therefore, no imputation and no analysis of missingness were required. Had missing data arisen, the mixed-effects framework would have accommodated observations missing at random. Given the small sample size, sensitivity analyses were not conducted, and this is acknowledged as a limitation. To account for within-subject variability in the crossover design, mixed-effects models were employed to test for a Time × Condition interaction. Potential carryover (period-by-sequence) was evaluated using the Sequence × Period interaction in a linear mixed-effects model. This approach avoided the use of pre–post difference scores, which have reduced sensitivity to condition effects.

## Results

The CONSORT flow diagram (Figure 2) illustrates participant enrolment, allocation to intervention groups, follow-up, and analysis, including reasons for exclusions and dropouts. See Supplementary Table S3 for CONSORT Checklist.

**Figure 2.**
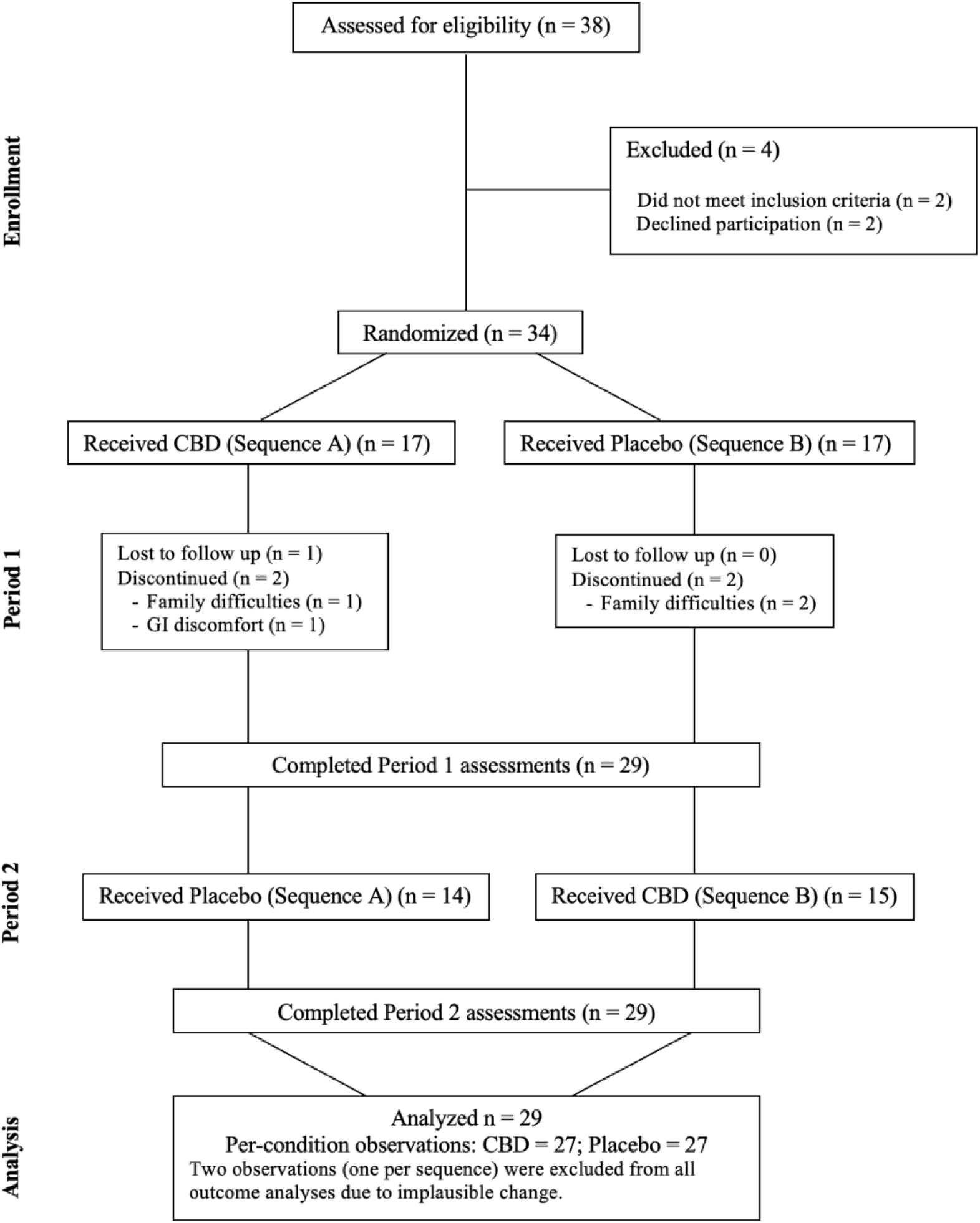
CONSORT flow diagram depicting enrolment, allocation, follow-up and analysis. Note: The two primary-outcome exclusions shown were from siblings in a single family (same caregiver informant). Decision to exclude was made prior to unblinding.

### Data completeness

Data completeness was 100% across measures. All participants who completed the trial provided both baseline and 12-week assessments, with no missing visits or item-level omissions.

### Baseline comparability and washout

Baseline scores at the start of each period were comparable (T0 vs T2) across all outcomes, consistent with an adequate 8-week washout (Supplementary Figures S1–S2).

### Primary Outcome

SRS-2: The Time by Intervention interaction indicated a greater reduction (i.e., improvement) in SRS-2 Total scores for CBD (mean difference = −17.0 points) compared to placebo (mean difference = −7.0 points); however, this interaction was not statistically significant (β = −11.15, SE = 7.19, *p* = .125) (Figure 3A).

**Figure 3.**
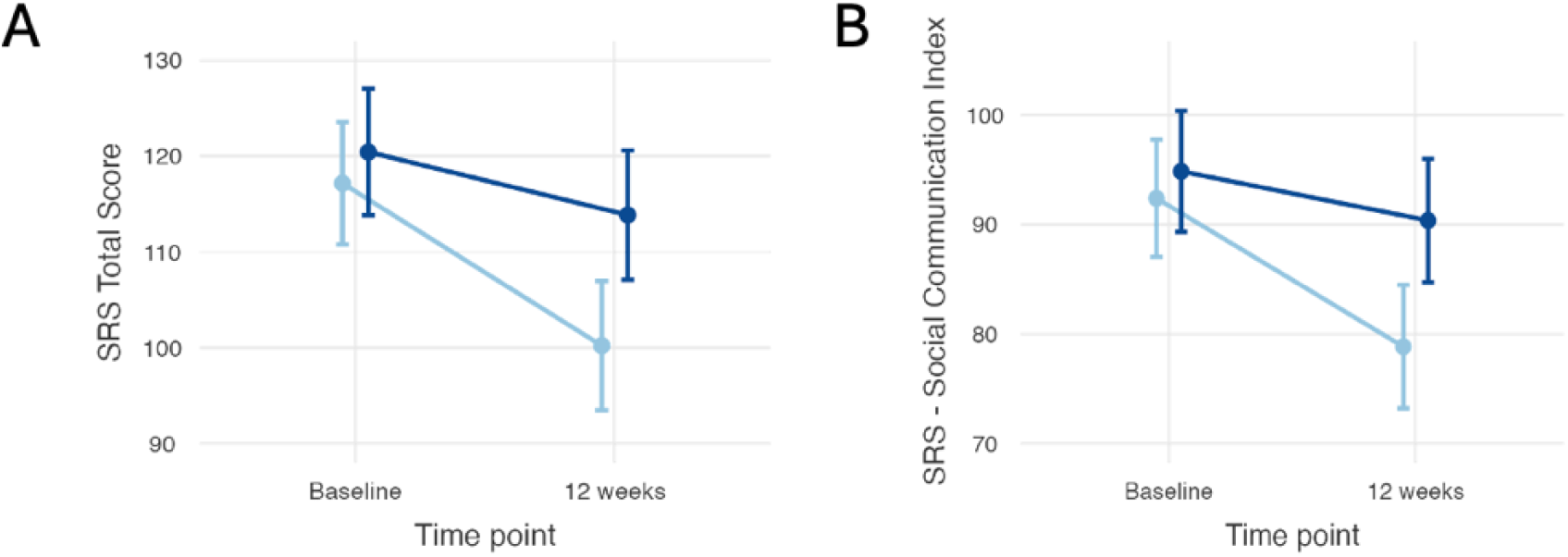
Time by Intervention effects of CBD (light blue) and placebo (dark blue) on (A) Social Responsiveness Scale (SRS−2) Total scores and (B) Social Communication Index composite scores from baseline to 12 weeks. Points are group means with standard-error (SE) bars. Lower scores indicate fewer social communication difficulties. N=27 participants.

For the SCI, the reduction in scores for CBD (mean difference = −13.8 points) compared to placebo (mean difference = −4.6 points) was not statistically significant (β = - 9.92, SE = 5.81, *p*_(adj)_ = .552) (Figure 3B). Change-from-baseline plots of the SRS-2 Total and SCI are provided in Figure 4a and Figure 4b, respectively. Greater reduction in scores following CBD compared to placebo were also observed for the SRS-2 subscales Social Awareness (CBD: −2.6 points, Placebo: −0.9 points; β = −1.77, SE = 1.030, *p*_(adj)_ = .540) and Social Motivation (CBD: −3.3 points, Placebo: −0.8 points; β = −2.48, SE = 1.265, *p*_(adj)_ = .318), although did not reach statistical significance. For Social Cognition (CBD: −2.8 points, Placebo: −1.3 points; β = −1.787, SE = 1.565, *p*_(adj)_ = 1.000), Social Communication (CBD: −5.1 points, Placebo: −1.7 points; β = −3.84, SE = 2.71, *p*_(adj)_ = .966), and Restricted Repetitive Behaviors (CBD: −2.9 points, Placebo: −2.1 points; β = −1.23, SE = 1.644, *p*_(adj)_ = 1.000) there were no intervention effects (Table 4).

**Figure 4.**
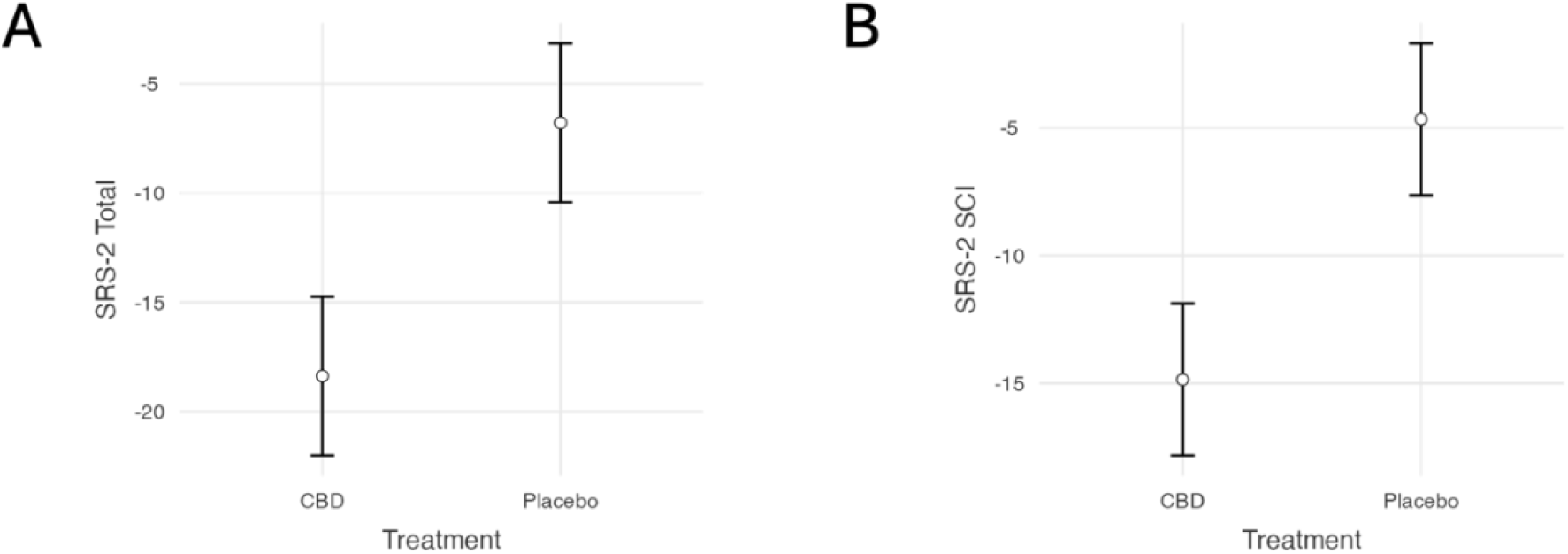
Change from baseline on SRS-2 outcomes by treatment. (A) SRS-2 Total T-score; (B) SRS-2 Social Communication Index. Points show the estimated mean change from baseline to 12 weeks for CBD and placebo across both crossover periods; vertical error bars indicate standard error (SE). Negative values indicate improvement (lower scores). N=27 participants.

### Secondary Outcomes

Analyses focus on the nine a priori secondary endpoints defined in the Methods.

#### Social Functioning

Three secondary measures probed social relating: PROMIS Social, Vineland Socialization, and DBC-2 Social Relating. For the PROMIS Social, an interaction effect with greater improvement following CBD intervention compared to placebo was observed but did not remain significant after Bonferroni correction (CBD: +1.7 points, Placebo: −0.2 points; β = 1.992, SE = 0.935, *p* = .036; p_(adj)_ = .072) (Figure 5A). There was no significant interaction effect observed for the Vineland-3 subscales Communication (CBD: +2.4 points, Placebo: +1.3 points; β = 1.82, SE = 2.40, *p* = .451; *p*_(adj)_ = 1.000) (Figure 5B) or Socialization (CBD: +4.4 points, Placebo: +2.4 points; β = 2.71, SE = 3.25, *p* = .406; *p*_(adj)_ = 1.000) (Figure 5C). Lastly, the DBC-2 Social Relating subscale showed a significant interaction effect (CBD: −2.55 points, Placebo: −0.29 points; β = −2.352, SE = 0.918, *p* = .012; *p*_(adj)_ = .024) (Figure 5D), suggesting a moderate impact of CBD intervention on social relating symptoms.

**Figure 5.**
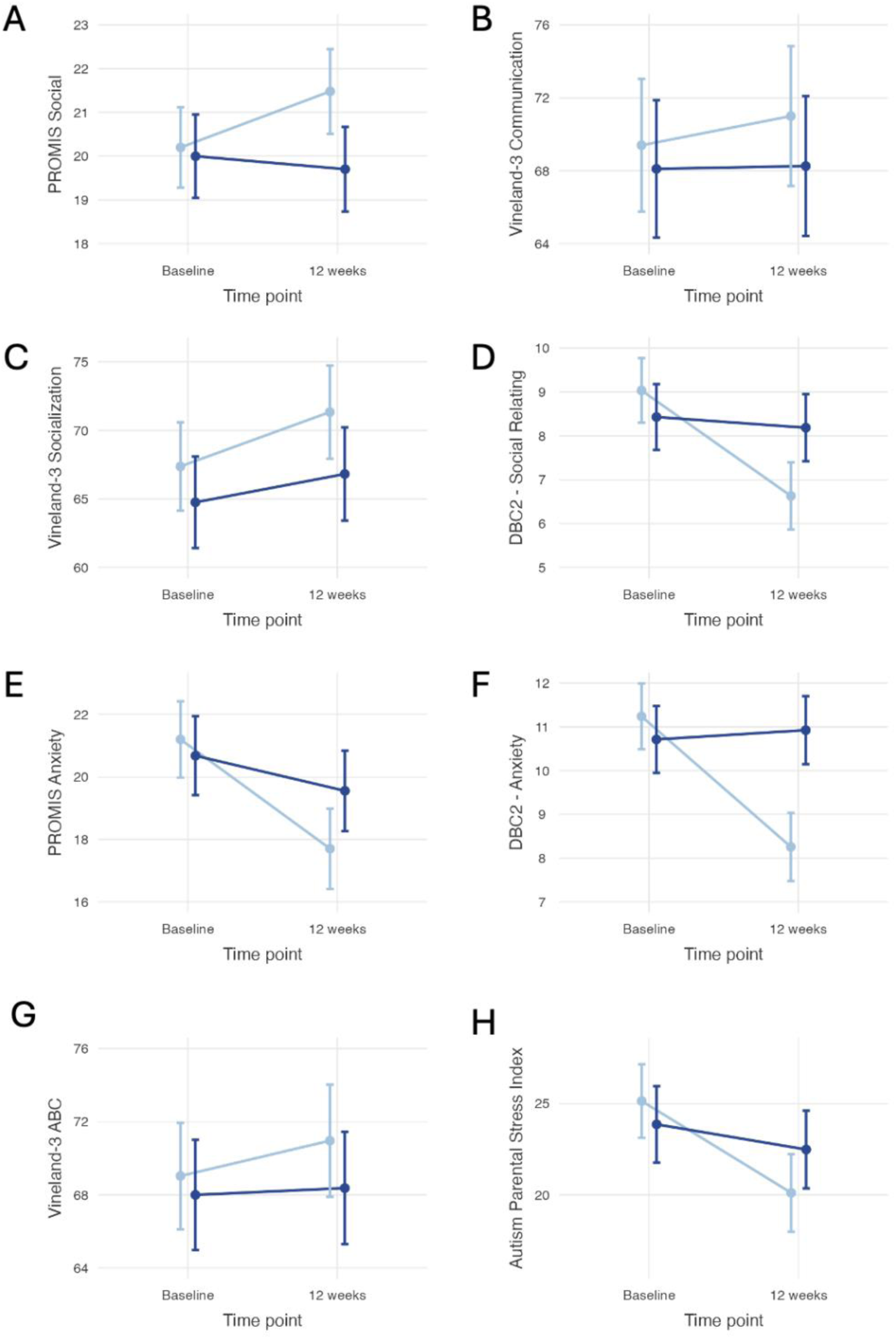
Time by Intervention effects of CBD (light blue) and placebo (dark blue) on (A) PROMIS Social, (B) Vineland-3 Communication, (C) Vineland-3 Socialization, (D) DBC-2- Social Relating, (E) PROMIS Anxiety, (F) DBC-2 - Anxiety, (G) Vineland-3 ABC, and (H) Autism Parental Stress Index. Points are group means with standard-error (SE) bars. Direction of improvement: Higher = improvement for Vineland-3 ABC, Vineland-3 Socialization, PROMIS Social; Lower = improvement for DBC-2 Social Relating, DBC-2 Anxiety, PROMIS Anxiety, Autism Parental Stress Index. N=27 participants.

#### Anxiety

The PROMIS Anxiety and the DBC-2 Anxiety subscales were used to measure changes in anxiety following CBD compared to placebo. For the PROMIS Anxiety subscale, there appeared to be a reduction in anxiety for CBD (mean difference = −4.1 points) compared to placebo (mean difference = −0.8 points); but the difference did not reach statistical significance (β = −3.119, SE = 1.660, *p* = .064; *p*_(adj)_ = .128) (Figure 5E). Conversely, a significant interaction effect for the Anxiety subscale from the DBC-2 was observed, indicating reduced anxiety symptoms following CBD (CBD: −3.14 points, Placebo: +0.2 points; β = −3.20, SE = 0.944, *p* = .001; *p*_(adj)_ = .002) (Figure 5F).

#### Adaptive Behaviour

There was no significant effect of CBD on Vineland-3 ABC scores compared to placebo (CBD: +2.4 points, Placebo: +1.0 points; β = 2.06, SE = 2.67, p = .443; *p*_(adj)_ = 1.000) (Figure 5G). Similarly, there were no effects of CBD on Vineland-3 subscale Living Skills (CBD: +1.0 points, Placebo: +0.1 points; β = 1.91, SE = 2.78, *p* = 0.493; *p*_(adj)_ = 1.000) (Table 5).

**Table 5.**
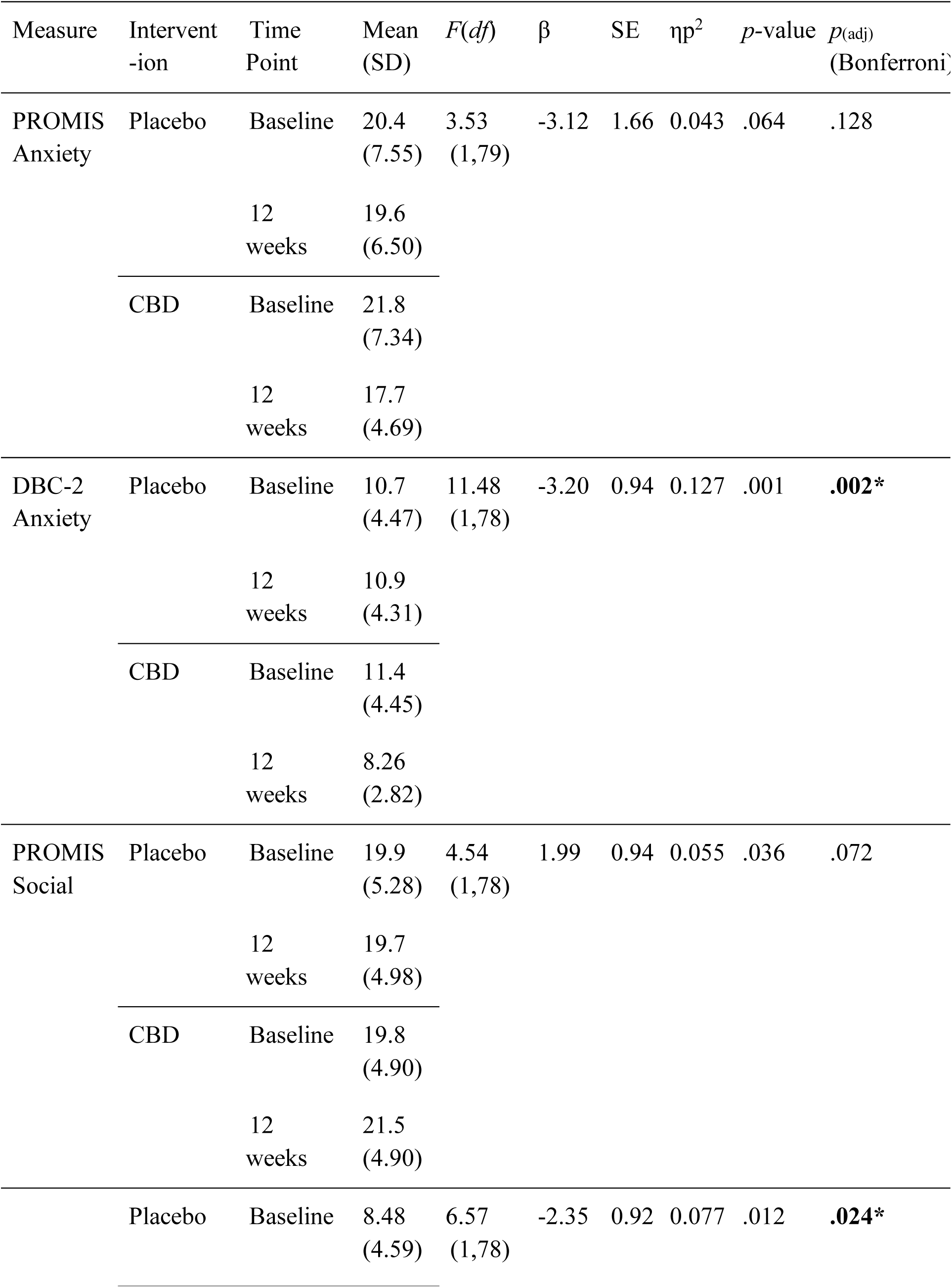
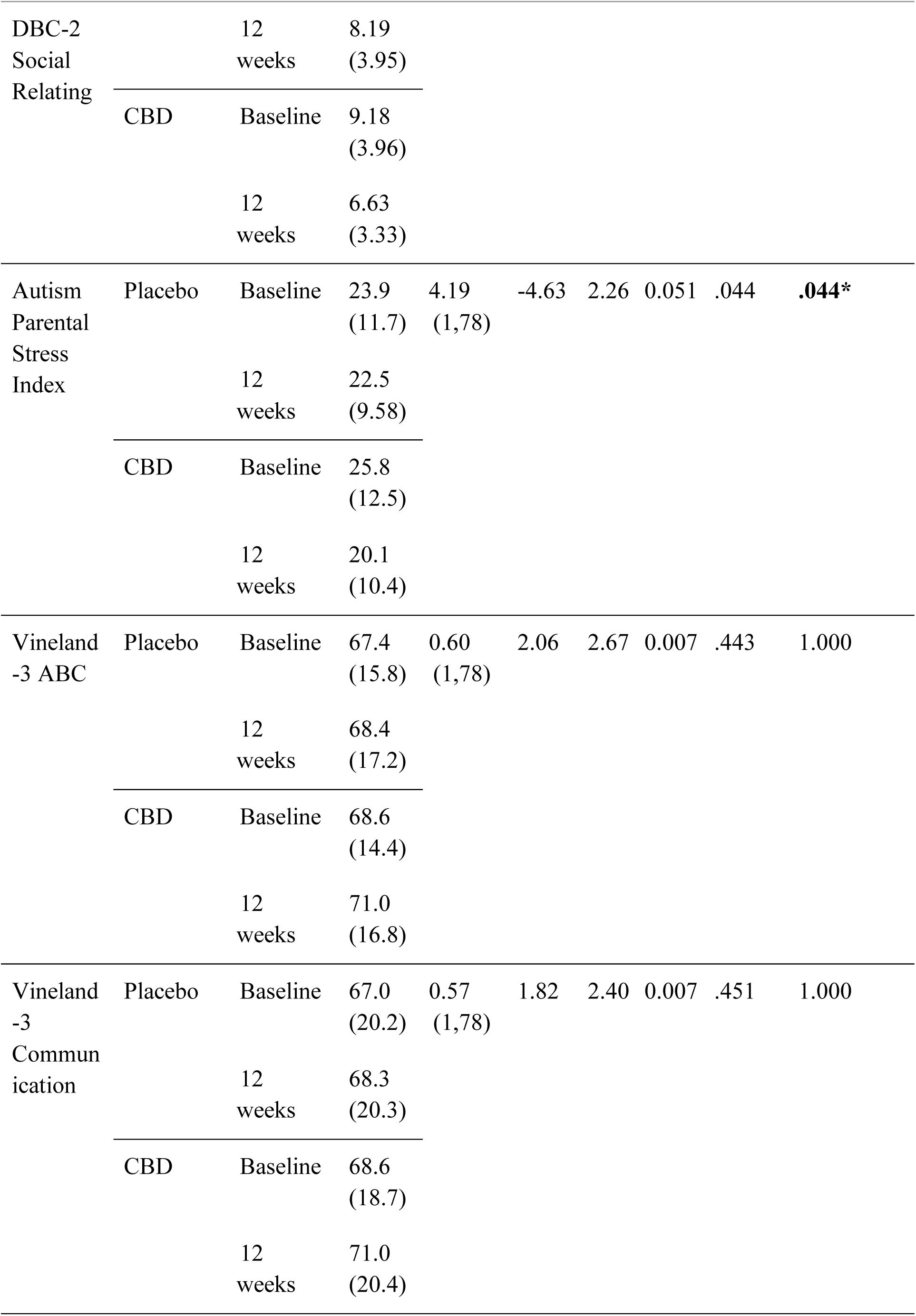
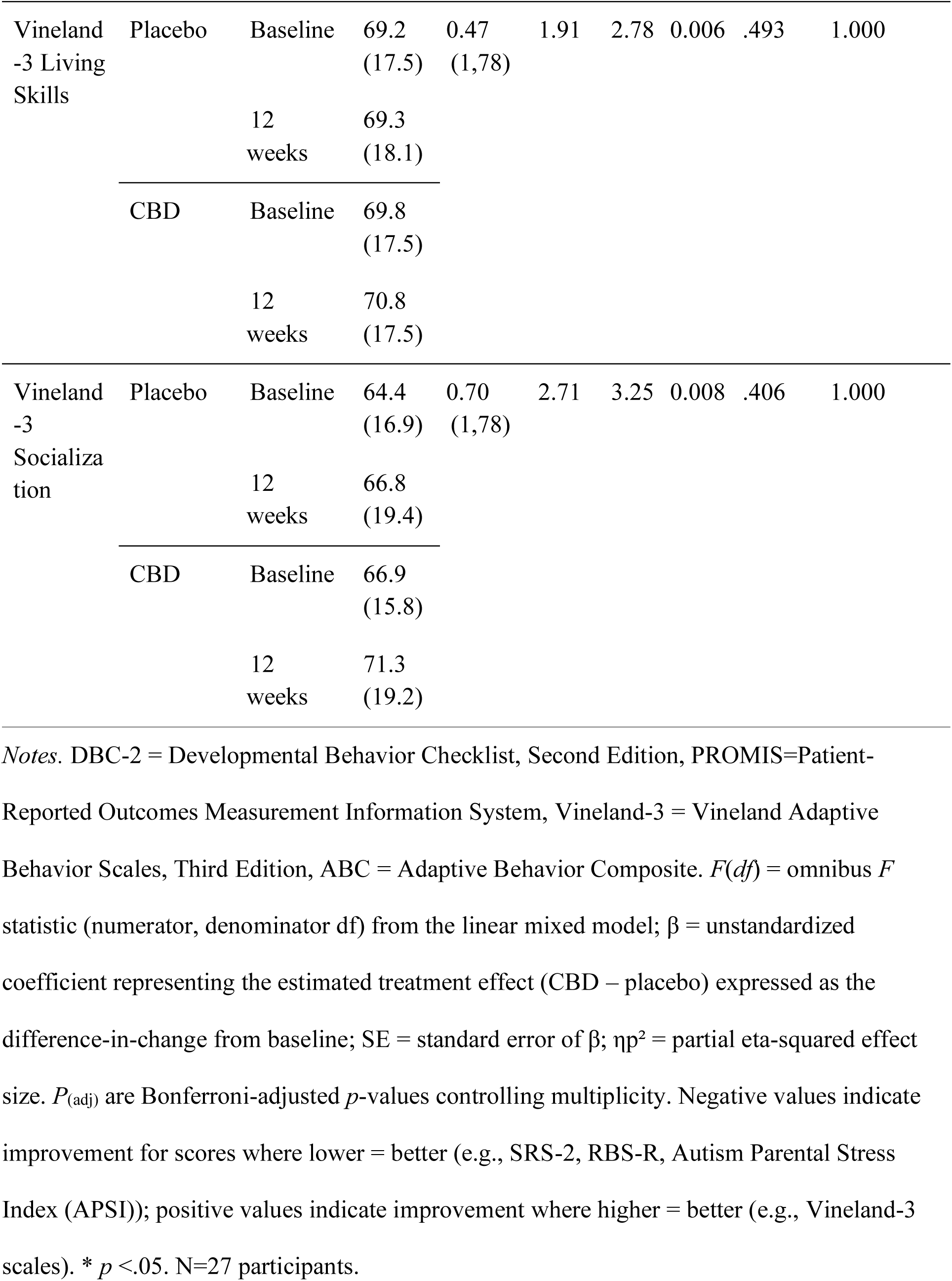
Summary of intervention effects on secondary outcome measures from baseline to 12-weeks.

| Measure | Intervention | Time Point | Mean (SD) | $F(df)$ | $\beta$ | SE | $\eta^2$ | $p$-value | $p_{(adj)}$ (Bonferroni) |
| --- | --- | --- | --- | --- | --- | --- | --- | --- | --- |
| PROMIS Anxiety | Placebo | Baseline | 20.4 (7.55) | 3.53 (1,79) | -3.12 | 1.66 | 0.043 | .064 | .128 |
|  |  | 12 weeks | 19.6 (6.50) |  |  |  |  |  |  |
|  | CBD | Baseline | 21.8 (7.34) |  |  |  |  |  |  |
|  |  | 12 weeks | 17.7 (4.69) |  |  |  |  |  |  |
| DBC-2 Anxiety | Placebo | Baseline | 10.7 (4.47) | 11.48 (1,78) | -3.20 | 0.94 | 0.127 | .001 | **.002\*** |
|  |  | 12 weeks | 10.9 (4.31) |  |  |  |  |  |  |
|  | CBD | Baseline | 11.4 (4.45) |  |  |  |  |  |  |
|  |  | 12 weeks | 8.26 (2.82) |  |  |  |  |  |  |
| PROMIS Social | Placebo | Baseline | 19.9 (5.28) | 4.54 (1,78) | 1.99 | 0.94 | 0.055 | .036 | .072 |
|  |  | 12 weeks | 19.7 (4.98) |  |  |  |  |  |  |
|  | CBD | Baseline | 19.8 (4.90) |  |  |  |  |  |  |
|  |  | 12 weeks | 21.5 (4.90) |  |  |  |  |  |  |
|  | Placebo | Baseline | 8.48 (4.59) | 6.57 (1,78) | -2.35 | 0.92 | 0.077 | .012 | **.024\*** |
| DBC-2<br>Social<br>Relating |  | 12<br>weeks | 8.19<br>(3.95) |  |  |  |  |  |  |
|  | CBD | Baseline | 9.18<br>(3.96) |  |  |  |  |  |  |
|  |  | 12<br>weeks | 6.63<br>(3.33) |  |  |  |  |  |  |
| Autism<br>Parental<br>Stress<br>Index | Placebo | Baseline | 23.9<br>(11.7) | 4.19<br>(1,78) | -4.63 | 2.26 | 0.051 | .044 | **.044\*** |
|  |  | 12<br>weeks | 22.5<br>(9.58) |  |  |  |  |  |  |
|  | CBD | Baseline | 25.8<br>(12.5) |  |  |  |  |  |  |
|  |  | 12<br>weeks | 20.1<br>(10.4) |  |  |  |  |  |  |
| Vineland<br>-3 ABC | Placebo | Baseline | 67.4<br>(15.8) | 0.60<br>(1,78) | 2.06 | 2.67 | 0.007 | .443 | 1.000 |
|  |  | 12<br>weeks | 68.4<br>(17.2) |  |  |  |  |  |  |
|  | CBD | Baseline | 68.6<br>(14.4) |  |  |  |  |  |  |
|  |  | 12<br>weeks | 71.0<br>(16.8) |  |  |  |  |  |  |
| Vineland<br>-3<br>Commun<br>ication | Placebo | Baseline | 67.0<br>(20.2) | 0.57<br>(1,78) | 1.82 | 2.40 | 0.007 | .451 | 1.000 |
|  |  | 12<br>weeks | 68.3<br>(20.3) |  |  |  |  |  |  |
|  | CBD | Baseline | 68.6<br>(18.7) |  |  |  |  |  |  |
|  |  | 12<br>weeks | 71.0<br>(20.4) |  |  |  |  |  |  |
| Vineland<br>-3 Living<br>Skills | Placebo | Baseline | 69.2<br>(17.5) | 0.47<br>(1,78) | 1.91 | 2.78 | 0.006 | .493 | 1.000 |
|  |  | 12<br>weeks | 69.3<br>(18.1) |  |  |  |  |  |  |
|  | CBD | Baseline | 69.8<br>(17.5) |  |  |  |  |  |  |
|  |  | 12<br>weeks | 70.8<br>(17.5) |  |  |  |  |  |  |
| Vineland<br>-3<br>Socializa<br>tion | Placebo | Baseline | 64.4<br>(16.9) | 0.70<br>(1,78) | 2.71 | 3.25 | 0.008 | .406 | 1.000 |
|  |  | 12<br>weeks | 66.8<br>(19.4) |  |  |  |  |  |  |
|  | CBD | Baseline | 66.9<br>(15.8) |  |  |  |  |  |  |
|  |  | 12<br>weeks | 71.3<br>(19.2) |  |  |  |  |  |  |

*Notes.* DBC-2 = Developmental Behavior Checklist, Second Edition, PROMIS=Patient-Reported Outcomes Measurement Information System, Vineland-3 = Vineland Adaptive Behavior Scales, Third Edition, ABC = Adaptive Behavior Composite. $F(df)$ = omnibus $F$ statistic (numerator, denominator df) from the linear mixed model; $\beta$ = unstandardized coefficient representing the estimated treatment effect (CBD – placebo) expressed as the difference-in-change from baseline; SE = standard error of $\beta$ ; $\eta^2$ = partial eta-squared effect size. $P_{(adj)}$ are Bonferroni-adjusted $p$ -values controlling multiplicity. Negative values indicate improvement for scores where lower = better (e.g., SRS-2, RBS-R, Autism Parental Stress Index (APSI)); positive values indicate improvement where higher = better (e.g., Vineland-3 scales). \* $p < .05$ . N=27 participants.

#### Parental Stress

There was a significant improvement in parental stress following the CBD intervention compared to placebo as measured by the APSI (CBD: −5.7 points, Placebo: −1.4 points; β = −4.6271, SE 2.26, *p* = .044, *p*_(adj)_ = .044) (Figure 5H). Change-from-baseline plots for PROMIS Social, Vineland-3 Communication, Vineland-3 Socialization, DBC-2 - Social Relating, PROMIS Anxiety, DBC-2 - Anxiety, Vineland-3 ABC, and Autism Parental Stress Index are shown in Figure 6.

**Figure 6.**
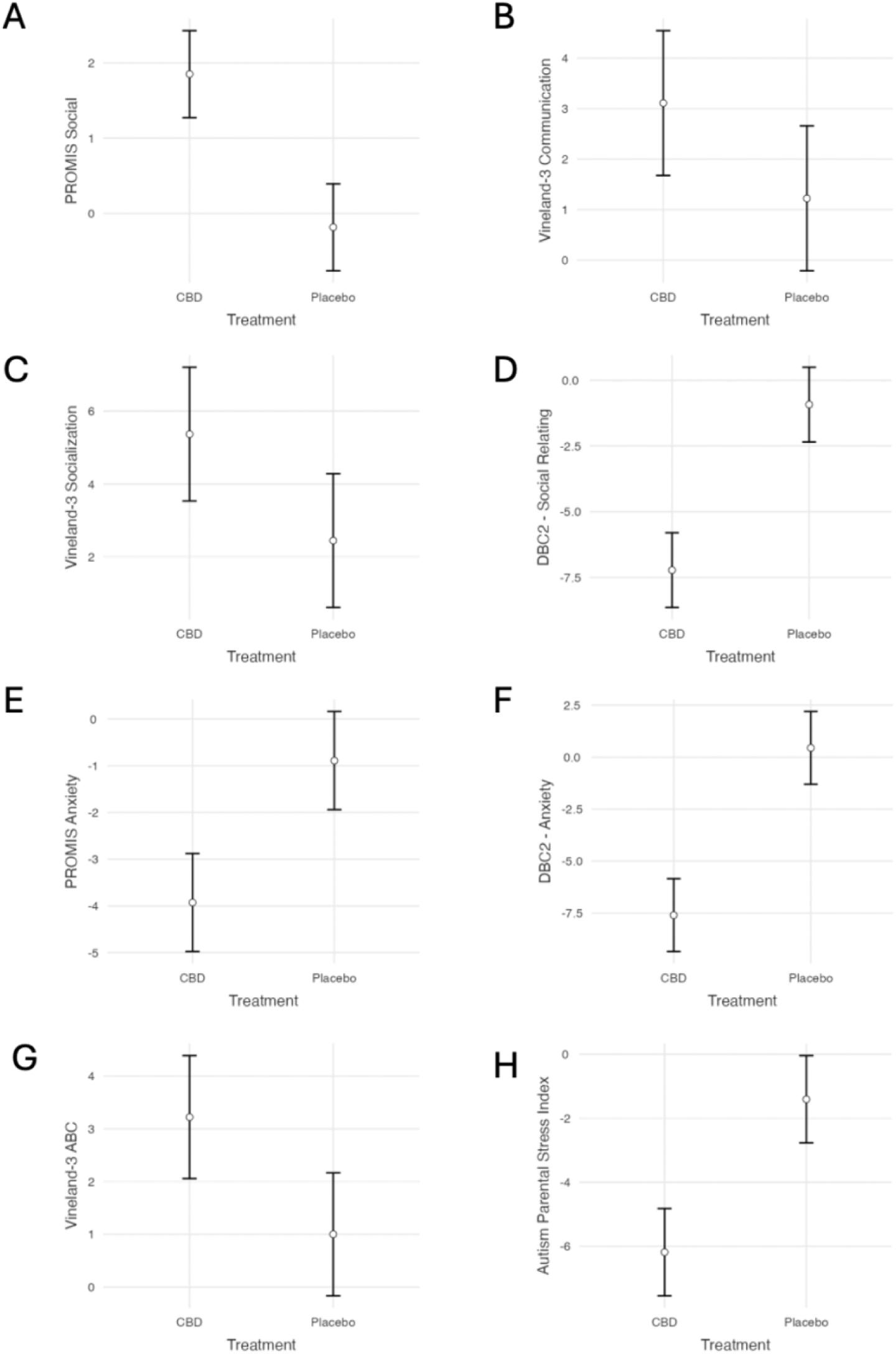
Change from baseline on secondary outcomes by treatment (Week 12 – Baseline). Panels: (A) PROMIS Social, (B) Vineland-3 Communication, (C) Vineland-3 Socialization, (D) DBC-2 - Social Relating, (E) PROMIS Anxiety, (F) DBC-2 - Anxiety, (G) Vineland-3 Adaptive Behavior Checklist (ABC), (H) Autism Parental Stress Index (APSI). Points show the estimated mean change across both crossover periods; vertical error bars show standard error (SE). Negative values indicate improvement on symptom scales where lower scores are better (e.g., APSI, PROMIS Anxiety, DBC-2 subscales); positive values indicate improvement where higher scores are better (e.g., Vineland-3 scales, PROMIS Social). Bonferroni-adjusted *p*-values are reported in the text. N=27 participants.

Intervention effects on the discussed outcome measures from baseline to 12-weeks are presented in Table 4 and Table 5. Exploratory analyses of the remaining secondary outcome measures are presented in the Supplementary Table S2.

Carryover was assessed via the Sequence × Period interaction. For the SRS outcomes, no interactions were significant (all *ps* > .10). Across PROMIS, DBC-2, Vineland, and APSI outcomes, no interactions reached statistical significance (all *p* > .057; smallest *p* = .056 for DBC-2 Anxiety). See Table 6 and Figure 7 for carryover results (statistics and model-estimated means, respectively).

**Figure 7.**
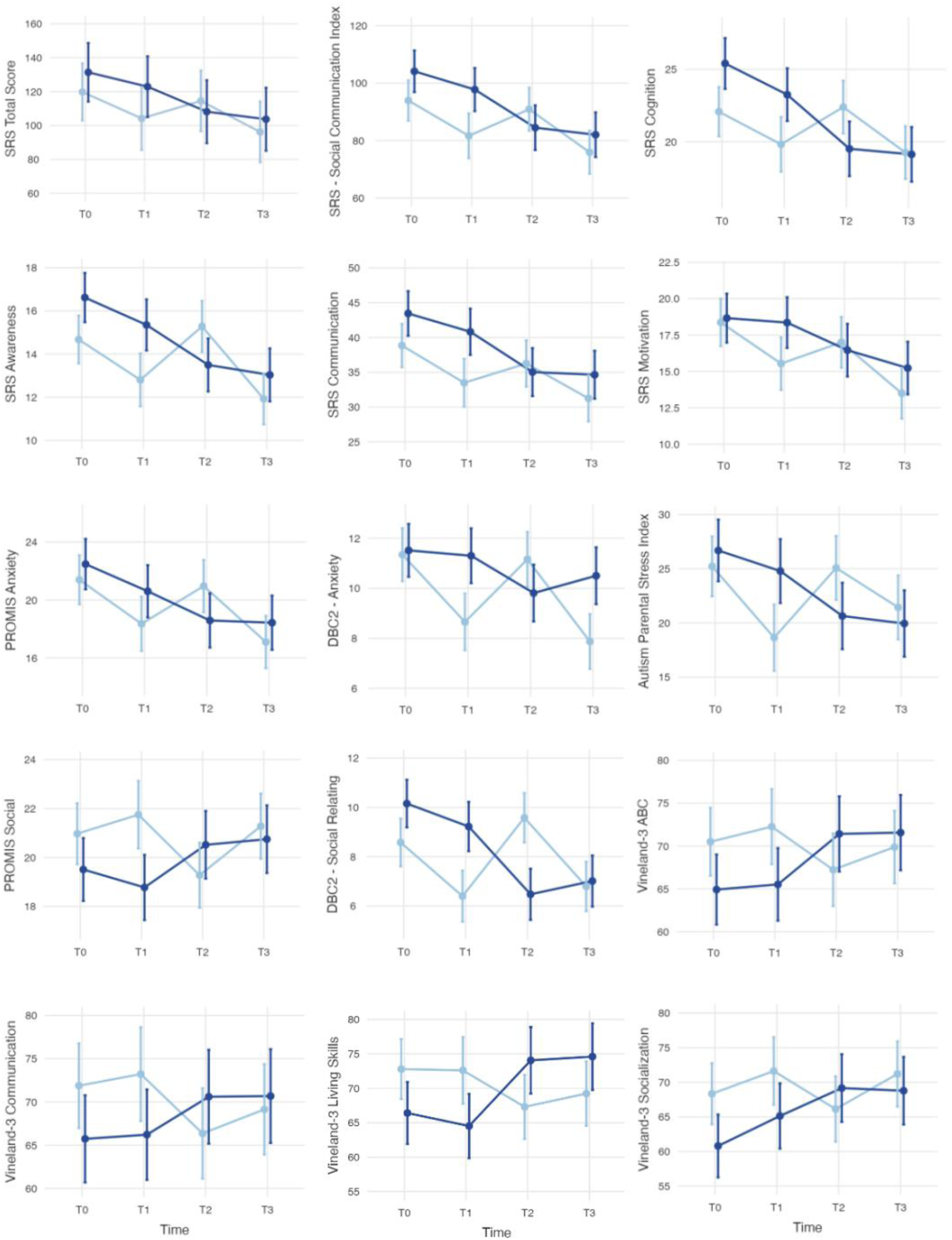
Model-estimated means for each outcome across the four assessment time points (T0–T3) by Intervention (CBD vs placebo) in the crossover design. Points are group means with standard-error (SE) bars. T0 and T1 correspond to baseline and 12 weeks in Period 1; T2 and T3 correspond to baseline and 12 weeks in Period 2, separated by an 8-week washout. Dark blue line = placebo; light blue line = CBD. *N* = 27 participants.

**Table 6.** Period-by-sequence (carryover) tests across outcomes at 12 weeks.

| Measure | Sequence × Period $F$ (1,49) | $p$-value | $\eta^2$ |
| --- | --- | --- | --- |
| **SRS Outcomes** |  |  |  |
| SRS-2 Total | 2.08 | .156 | .041 |
| SRS-2 – SCI | 2.16 | .148 | .042 |
| SRS-2 – Social Awareness | 2.41 | .127 | .047 |
| SRS-2 – Social Cognition | 0.88 | .353 | .018 |
| SRS-2 – Social Communication | 2.56 | .116 | .050 |
| SRS-2 – Social Motivation | 1.53 | .222 | .030 |
| SRS-2 – RRB | 1.04 | .312 | .021 |
| **Secondary Outcomes** |  |  |  |
| PROMIS Anxiety | 1.32 | .256 | .026 |
| DBC-2 – Anxiety | 3.79 | .057 | .072 |
| PROMIS Social | 1.73 | .194 | .034 |
| DBC-2 – Social Relating | 2.56 | .116 | .050 |
| Autism Parental Stress Index | 0.73 | .396 | .015 |
| Vineland-3 ABC | 0.30 | .588 | .006 |
| Vineland-3 Communication | 0.25 | .618 | .005 |

| Measure | Sequence × Period $F(1,49)$ | $p$-value | $\eta^2$ |
| --- | --- | --- | --- |
| Vineland-3 Living Skills | 0.08 | .780 | .002 |
| Vineland-3 Socialization | 0.70 | .407 | .014 |

*Notes.* SRS-2 = Social Responsiveness Scale, Second Edition; SCI = Social Communication Index; RRB = Restricted and Repetitive Behaviors; PROMIS = Patient-Reported Outcomes Measurement Information System; DBC-2 = Developmental Behavior Checklist, Second Edition; Vineland-3 = Vineland Adaptive Behavior Scales, Third Edition; ABC = Adaptive Behavior Composite. “Sequence × Period” denotes the period-by-sequence (carryover) test from the linear mixed model (fixed effects: sequence, period, sequence × period; random intercept for subject ID; Sequence A = CBD-Placebo, Sequence B = Placebo-CBD). $F(1,49)$ = omnibus $F$ statistic with numerator and denominator degrees of freedom; $\eta^2$ = partial eta-squared. \* $p < .05$ . $N = 27$ participants.

### Adverse Events

Parents completed a daily Treatment Log that specifically probed for adverse events and general health/wellbeing, ensuring regular assessment of potential side effects throughout the trial. During the trial, there were two adverse events recorded. One participant experienced abdominal discomfort on day 4 of CBD intervention, as reported by their parent, leading to the discontinuation of intervention after 7-days and withdrawal from the trial (Figure 2). This participant had pre-existing gastrointestinal issues, which may have contributed to the reported discomfort. Another participant also reported gastrointestinal discomfort during the CBD condition, but this subsided after one week and the participant remained in the trial. Medical consultations were provided to the parents by a licensed medical practitioner, offering guidance on managing symptoms and assessing the suitability of continued participation in the study. No other adverse events were reported throughout the trial duration, indicating a generally well-tolerated intervention.

## Discussion

This randomized, double-blind, placebo-controlled, crossover trial is among the first to investigate the effects of oral CBD oil on social relating in autistic children, with anxiety and parental stress as secondary outcomes. The trial addresses a critical gap by evaluating chronic CBD administration in autistic children, an area with few RCTs systematically assessing long-term effects, despite the increase in off-label prescribing of CBD in this population.

There was no significant change in the primary outcome measure of social relating (SRS-2) following 12 weeks of CBD oil compared to placebo oil. However, changes in the SRS-2 Total score and Social Communication Index were directionally consistent with potential improvement, and SRS-2 Total scores decreased by an average of 17 points following CBD oil compared with a 7-point reduction following placebo oil. The SRS-2 manual states that a 10-point raw-score shift corresponds to one severity-band reduction, indicating that a 17-point decrease may be clinically meaningful if confirmed by larger trials. The direction of our findings align with a previous placebo-controlled, double-blind, crossover study in autistic children and young adults, that reported a 14.9-point decrease on the SRS (secondary outcome) following 12-weeks of CBD:THC 20:1 whole-plant extract intervention (1 to ∼7.5-10 mg/kg/day, n=34) (Aran et al., 2021). However, given the pilot sample size and phenotypic heterogeneity (e.g., cognitive/language variability), the present study was likely underpowered, thus these observations remain exploratory and hypothesis-generating rather than evidencing efficacy.

The present study employed both total and subscale scores from the SRS-2 as outcome measures to capture global social responsiveness as well as domain-specific effects, thereby providing a comprehensive evaluation of the impact of CBD on social functioning. Given that 11 participants had intellectual disability, it is important to note that certain assessment measures may have been better suited for their evaluation than others. For instance, individuals who were non-verbal or faced sensory challenges might not have been adequately captured by measures emphasizing higher levels of communication abilities such as the SRS-2. The effect of measure is evidenced by the significant effects of CBD on other social functioning measures, including the DBC-2 Social Relating subscale, which indicated a moderate positive effect of CBD intervention on social relating symptoms. The 12-week CBD intervention also significantly reduced DBC-2 Anxiety subscale, which demonstrates good convergence with interview-based anxiety diagnoses in autistic young people (Halvorsen et al., 2025). These outcomes suggest a potential interplay between social anxiety and social relating, whereby reducing anxiety may improve social interactions (or vice-versa) (Bergamaschi et al., 2011), although CBD could also act on both domains simultaneously (Crippa et al., 2011; Shannon, 2019; Silva Junior et al., 2024). The observed improvements highlight the need for further research in this area, particularly considering the limited pharmacological options available for addressing social difficulties and anxiety in autism (Warren et al., 2011). Although subscale-specific thresholds are limited, recent work indicates that a change of 3-6 points on the DBC-2 Total Behaviour Problems Score is considered clinically important (Sutherland et al., 2025). Our Anxiety subscale effect (reduction of 3.2 points) is within this range, suggesting possible clinical meaningfulness. In line with a multicenter phase-II RCT of purified CBD oral solution (10 mg/kg/day) (GW Research Ltd, 2024), we also observed no significant effects on Vineland-3 domains.

Parents/guardians showed an improvement in parental stress scores, measured by the APSI, following the 12-week CBD intervention in their children. The 4.6 point reduction observed in this trial can be interpreted as a small-to-moderate improvement when considered against distribution-based benchmarks for meaningful change (Norman et al., 2003). Original validation of the APSI demonstrated strong reliability and demonstrates feasibility (Silva & Schalock, 2012). More recent work in newly diagnosed autistic children reports good internal consistency for the APSI, with and higher scores associated with poorer family quality of life (Papadopoulos et al., 2023). Parenting an autistic child has been associated with greater parenting stress compared with parenting a neurotypical child, and improvements in parental well-being have been linked to enhanced outcomes for autistic children, including reduced behavioral difficulties (Hayes & Watson, 2013). This finding raises questions regarding the attribution of child improvement following CBD, parenting factors, or a combination of both. It is also possible that parents willing to enroll their child in an RCT might differ in certain characteristics, such as being more proactive in seeking interventions or having different stress-coping strategies, compared to those who do not participate. This significant finding suggests there is value in probing both child and parental well-being in autism-related clinical trials.

This trial demonstrates that CBD may be a promising intervention for social relating in autism due to its favourable safety profile and lack of intoxicating effects, making it a plausible candidate for further investigation in pediatric populations. It is also important to note that participants continued their previously prescribed medications for the duration of the trial. Coupled with the overall low number of reported adverse events (n=2), this suggests that 10 mg/kg/day of CBD could be taken safely alongside existing medications in these children. The two reported adverse events were gastrointestinal discomfort during the CBD condition, with one child withdrawing from the trial before titrating to 10 mg/kg/day. Given the oil-based dosage format used in the present study, gastrointestinal discomfort is not an unexpected adverse event (Bar-Lev Schleider et al., 2019; Efron et al., 2021; Fleury-Teixeira et al., 2019). However, this does prompt speculation on whether the dosage of CBD administered may have been too high for some participants, and whether a lower dose, or higher concentration of CBD to oil (e.g., 200 mg/mL), could potentially yield similar beneficial outcomes with fewer side effects. Further investigation into dosage optimization is warranted to address these concerns and enhance tolerability in future trials.

Overall, the present trial’s intervention and outcomes both align and contrast with previous studies. Aran and colleagues randomized 150 participants to 2 of 3 interventions: whole-plant extract (CBD:THC 20:1), purified CBD+THC preparation at the same ratio, or placebo, each for 12 weeks in a double-blind, crossover design. The SRS-2 Total (secondary outcome) improved more with the whole-plant extract than placebo (−14.9 vs −3.6 points), whereas the primary outcome measures, which assessed disruptive behavior, showed mixed findings. CBD was titrated from 1 to 5-10 mg/kg/day (THC 0.05 mg/kg/day), averaging ∼5.7 mg/kg/day (Aran et al., 2021). In contrast, our trial evaluated a ‘broad-spectrum’ CBD + terpenes oil with negligible THC, prioritized social relating (SRS-2) as the primary outcome, and titrated to 10 mg/kg/day CBD oil, which may account for differences in findings. Silva Junior and colleagues reported improvements in social interaction, anxiety, psychomotor agitation, meal frequency, and concentration over 12 weeks using a CBD:THC 9:1 extract with dosing recorded as number of drops. Their methodology limits comparability with the present study due to its parallel-arm design, lower CBD:THC ratio, lack of precise dose reporting, and use of different outcome measures (Autism Treatment Evaluation Checklist and semi-structured interviews) (Silva Junior et al., 2024). Overall, differences in formulation/THC, dosing, and outcome measures likely explain discrepant findings and support eventual standardization of pediatric RCTs targeting core social outcomes (Parrella et al., 2023).

The present trial investigated Medigrowth CBD100 (<1 mg/mL THC), which relies on CBD-mediated mechanisms rather than THC-driven CB_1_ agonism. Existing RCTs have administered distinct cannabinoid formulations, including a 20:1 CBD:THC whole-plant extract (Aran et al., 2021), a 98% purified CBD isolate (Efron et al., 2021), and a 9:1 CBD:THC extract (Silva Junior et al., 2024). As noted above, the present formulation choice is consistent with the selective improvements observed on secondary measures, whereas larger effects have been reported with lower CBD:THC ratio whole-plant extracts (Aran et al., 2021). Use of products with a lower CBD ratio in children raises clinical and regulatory considerations, as 20:1 and 9:1 CBD:THC formulations are classified as Schedule 8 Controlled Drugs in Australia, requiring GPs to have specialist endorsement before seeking government approval to prescribe (Therapeutic Goods Administration (TGA), 2025). Medigrowth CBD100 also retains terpenes, with effects expected to arise from FAAH inhibition, partial 5-HT_1_A agonism, and positive allosteric modulation of GABA_A_ receptors (Bakas et al., 2017; Bisogno et al., 2001; Campos & Guimarães, 2008). Thus, specific terpenes, such as Nerolidol, α-Bisabolol, d-Limonene, β-Caryophyllene, and α-Humulene, may have contributed to the therapeutic effects observed and would be consistent with the proposed synergistic interactions between cannabinoids and terpenes (Russo, 2011).

The premise of this research stemmed from the growing recognition of the involvement of the ECS in social relating, specifically in autism, suggesting a potential role for CBD in modulating social functioning for autistic children (Wei et al., 2015; Zamberletti et al., 2017). Evidence from preclinical and human studies suggests that CBD can enhance endocannabinoid tone by limiting anandamide (AEA) hydrolysis and may modulate signaling at several G protein-coupled receptors (GPCRs) (Karhson et al., 2016; Wei et al., 2016). Preclinical studies have reported that CBD directly inhibits fatty acid amide hydrolase (FAAH) (Bisogno et al., 2001), while human *in vitro* data indicate CBD binds fatty-acid binding proteins (FABPs) that transport anandamide (AEA) to FAAH, limiting AEA hydrolysis (Elmes et al., 2015). At similar concentrations, CBD has been shown to modulate 5-hydroxytryptamine (serotonin) 1A (5-HT₁A), gamma-aminobutyric acid (GABA)_A_, and transient receptor potential vanilloid 1 (TRPV1) receptors (Bakas et al., 2017; Campos & Guimarães, 2008; Iseger & Bossong, 2015). These mechanisms may influence circuits relevant to social-reward processing and reduce amygdala-driven anxiety (Wei et al., 2016; Zamberletti et al., 2017). In addition, magnetic resonance spectroscopy (MRS) data has shown acute shifts in cortical glutamate and GABA after a single CBD dose in autistic adults, suggesting that re-balancing excitatory-inhibitory neurotransmission may also attenuate hyperactivity and related behaviors, although evidence remains preliminary (Pretzsch, Freyberg, et al., 2019). Serotonergic, adenosinergic, GABAergic, and endocannabinoid pathways could also underlie the improvements in hyperactivity, sleep, and anxiety reported in some smaller, preliminary studies of CBD in autism (Aran et al., 2019; Barchel et al., 2019); however, findings remain mixed and variable (Parrella et al., 2023).

There were several limitations that require consideration when interpreting the results of this trial. First, the pilot-sized sample and single-center design may constrain the generalizability of the reported findings. Second, due to relatively small sample size, stratification was not feasible, although it would have benefited the heterogeneity observed across participants, particularly regarding cognitive levels and language abilities. Third, although all participants had documented clinical diagnoses of ASD (per DSM-5 criteria) confirmed by external clinicians, we did not conduct study-administered diagnostic re-verification using standardized observational tools (e.g., ADOS-2/CARS-2). Reliance on prior specialist diagnosis may have introduced diagnostic heterogeneity. Future larger trials should incorporate standardized re-verification and pre-specified stratification (e.g., by language/cognitive level) to reduce heterogeneity.

Two parents anecdotally reported differences in oil consistency between the CBD and placebo formulations. This difference may have increased the risk of unblinding, particularly in families with siblings assigned to different conditions. We did not administer a post-study blinding questionnaire; therefore, blinding integrity could not be verified. Thus, future studies should standardise formulations, assess their impact on outcome measures, and include a formal blinding-integrity check. Although the Treatment Log offered prompts and an open-text field, we did not administer a validated symptom checklist. Consequently, very mild or transient events may not have been reported, potentially underestimating the true adverse event frequency. Despite limitations concerning statistical power, cohort heterogeneity, oil formulation, diagnostic verification, and adverse event assessment, the efficient recruitment observed in this pilot study suggests the feasibility of conducting larger investigations to explore the potential benefits of CBD in managing challenges associated with autism.

Future research endeavors should aim to conduct larger-scale, multi-site trials. By involving multiple research sites, trials can recruit larger and more diverse participant pools, improve the representativeness of findings, and increase statistical power. In larger trials containing clinical heterogeneity among participants, stratification could provide insights into intervention mechanisms by identifying specific characteristics within subgroups. They could also incorporate additional interim assessments to characterize the trajectory of change across treatment. Additionally, the anxiolytic and sleep effects of CBD have been identified as exhibiting an inverted U-shape dose-response curve, highlighting the complexity and critical nature of dosage and administration methods in clinical trials involving this compound (Narayan et al., 2022). Tailoring dosages to individual responsivity could also lead to more precise and effective provision of CBD. Given the lack of effective pharmacological therapeutics for social relating in autism, without side effects (Nurmi et al., 2013; Sharma & Shaw, 2012), there is a critical need to develop novel targeted interventions.

## Conclusion

This RCT undertook a symptom-specific approach to investigating the effects of CBD on social relating in autistic children. By combining CBD administration with behavioral testing specifically targeting social relating, this study offers a comprehensive exploration of the therapeutic effects of CBD in a population with limited pharmacological therapeutic options. The preliminary findings contribute to the current understanding regarding novel therapeutics with acceptable safety profiles to support the social difficulties experienced by autistic children. The results of this pilot trial suggest that chronic CBD intervention may improve social relating, anxiety and parental stress levels, with minimal side effects. These preliminary findings call for larger, multi-site trials to elucidate the therapeutic efficacy of CBD in treating social challenges associated with autism and provide direction on which outcome measures should be employed. Overall, this trial serves as a critical step towards enhancing our understanding of the neurobiological underpinnings of autism and fostering the development of targeted therapeutics that can improve the quality of life for autistic people.

## Data Availability

All data produced in the present study are available upon reasonable request to the authors

## Ethics Approval Statement

This study received ethics approval from the Deakin University Human Research Ethics Committee (DUHREC 2020-071) prior to enrolment of the first participant.

## Patient Consent Statement

Written informed consent was obtained from the parents or guardians of all participants. Assent was obtained from participants when appropriate.

## Clinical Trial Registration

This trial was prospectively registered with the Australian New Zealand Clinical Trials Registry (ACTRN12622000437763).

## Data Availability Statement

The datasets generated and analyzed during the current study are available from the corresponding author upon reasonable request.

## Permission to Reproduce Material

No third-party copyrighted material was reproduced in this manuscript.

## References

1. American Psychiatric Association. (2022). Diagnostic and Statistical Manual of Mental Disorders (5th ed., text rev.). American Psychiatric Association Publishing.

2. Aran, A., Eylon, M., Harel, M., Polianski, L., Nemirovski, A., Tepper, S., Schnapp, A., Cassuto, H., Wattad, N., & Tam, J. (2019). Lower circulating endocannabinoid levels in children with autism spectrum disorder. Molecular Autism, 10(1), 2. 10.1186/s13229-019-0256-6

3. Aran, A., Harel, M., Cassuto, H., Polyansky, L., Schnapp, A., Wattad, N., Shmueli, D., Golan, D., & Castellanos, F. X. (2021). Cannabinoid treatment for autism: A proof-of-concept randomized trial. Molecular Autism, 12(1), 6. 10.1186/s13229-021-00420-2

4. Bakas, T., Van Nieuwenhuijzen, P. S., Devenish, S. O., McGregor, I. S., Arnold, J. C., & Chebib, M. (2017). The direct actions of cannabidiol and 2-arachidonoyl glycerol at GABA A receptors. Pharmacological Research, 119, 358–370. 10.1016/j.phrs.2017.02.022

5. Baldes, A., May, T., Brignell, A., & Williams, K. (2024). Patterns of Psychotropic Prescribing Practices in Autistic Children and Adolescents: An Australian Perspective of Two Cohorts Five Years Apart. Child Psychiatry & Human Development. 10.1007/s10578-024-01710-5

6. Barchel, D., Stolar, O., De-Haan, T., Ziv-Baran, T., Saban, N., Fuchs, D. O., Koren, G., & Berkovitch, M. (2019). Oral Cannabidiol Use in Children With Autism Spectrum Disorder to Treat Related Symptoms and Co-morbidities. Frontiers in Pharmacology, 9, 1521.

7. Bar-Lev Schleider, L., Mechoulam, R., Saban, N., Meiri, G., & Novack, V. (2019). Real life Experience of Medical Cannabis Treatment in Autism: Analysis of Safety and Efficacy. Scientific Reports, 9(1), 200.

8. Bisogno, T., Hanuš, L., De Petrocellis, L., Tchilibon, S., Ponde, D. E., Brandi, I., Moriello, A. S., Davis, J. B., Mechoulam, R., & Di Marzo, V. (2001). Molecular targets for cannabidiol and its synthetic analogues: Effect on vanilloid VR1 receptors and on the cellular uptake and enzymatic hydrolysis of anandamide: Cannabidiol, VR1 receptors and anandamide inactivation. British Journal of Pharmacology, 134(4), 845–852. 10.1038/sj.bjp.0704327

9. Bodfish, J. W., Symons, F. J., Parker, D. E., & Lewis, M. H. (2000). Varieties of repetitive behavior in autism: Comparisons to mental retardation. Journal of Autism and Developmental Disorders, 30(3), 237–243. 10.1023/A:1005596502855

10. Buchwald, K., Shepherd, D., Siegert, R. J., Vignes, M., & Landon, J. (2025). Factors predicting parenting stress in the autism spectrum disorder context: A network analysis approach. PLOS One, 20(4), e0319036. 10.1371/journal.pone.0319036

11. Campos, A. C., & Guimarães, F. S. (2008). Involvement of 5HT1A receptors in the anxiolytic-like effects of cannabidiol injected into the dorsolateral periaqueductal gray of rats. Psychopharmacology, 199(2), 223–230. 10.1007/s00213-008-1168-x

12. Chin, R., Gil-Nagel, A., Mitchell, W., Patel, A. D., Perry, M. S., Weinstock, A., Checketts, D., & Dunayevich, E. (2020). Long-term safety and efficacy of Cannabidiol (CBD) treatment in Lennox Gastaut syndrome: Results overall and for patients completing 1-3 years of an open-label extension (GWPCARE5). European Journal of Neurology, 27, 24.

13. Cummins, R. A., & Lau, A. L. D. (2005). Personal Wellbeing Index – School Children (PWI-SC): Manual. Australian Centre on Quality of Life, Deakin University.

14. Devinsky, O., Patel, A. D., Thiele, E. A., Wong, M. H., Appleton, R., Harden, C. L., Greenwood, S., Morrison, G., Sommerville, K., & On behalf of the GWPCARE1 Part A Study Group. (2018). Randomized, dose-ranging safety trial of cannabidiol in Dravet syndrome. Neurology, 90(14), e1204–e1211. 10.1212/WNL.0000000000005254

15. Efron, D., Freeman, J. L., Cranswick, N., Payne, J. M., Mulraney, M., Prakash, C., Lee, K. J., Taylor, K., & Williams, K. (2021). A pilot randomised placebo-controlled trial of cannabidiol to reduce severe behavioural problems in children and adolescents with intellectual disability. British Journal of Clinical Pharmacology, 87(2), 436–446.

16. Einfeld, S. L., & Tonge, B. J. (2002). Manual for the Developmental Behaviour Checklist (Second Edition) (DBC-2). Monash University Centre for Developmental Psychiatry & Psychology; University of New South Wales School of Psychiatry.

17. Elmes, M. W., Kaczocha, M., Berger, W. T., Leung, K., Ralph, B. P., Wang, L., Sweeney, J. M., Miyauchi, J. T., Tsirka, S. E., Ojima, I., & Deutsch, D. G. (2015). Fatty Acid-binding Proteins (FABPs) Are Intracellular Carriers for Δ9-Tetrahydrocannabinol (THC) and Cannabidiol (CBD). Journal of Biological Chemistry, 290(14), 8711–8721. 10.1074/jbc.M114.618447

18. Faul, F., Erdfelder, E., Lang, A.-G., & Buchner, A. (2007). G*Power 3: A flexible statistical power analysis program for the social, behavioral, and biomedical sciences. Behavior Research Methods, 39(2), 175–191. 10.3758/BF03193146

19. Fleury-Teixeira, P., Caixeta, F. V., Ramires da Silva, L., Cruz, ro, Brasil-Neto, J. P., & Malcher-Lopes, R. (2019). Effects of CBD-Enriched Cannabis sativa Extract on Autism Spectrum Disorder Symptoms: An Observational Study of 18 Participants Undergoing Compassionate Use. Frontiers in Neurology, 10, 1145.

20. Gioia, G. A., Isquith, P. K., Guy, S. C., & Kenworthy, L. (2015). Behavior Rating Inventory of Executive Function, Second Edition (BRIEF). Psychological Assessment Resources.

21. GW Research Ltd. (2024). *An exploratory, Phase 2, randomized, double-blind, placebo-controlled trial to investigate the safety and efficacy of cannabidiol oral solution (GWP42003-P; CBD-OS) in children and adolescents with Autism Spectrum Disorder* [EudraCT No. 2020-002819-21]. EU Clinical Trials Register. https://www.clinicaltrialsregister.eu/ctr-search/trial/2020-002819-21/results

22. Halvorsen, M. B., Heiervang, E. R., Mathiassen, B., Aman, M. G., Kaiser, S., & Helverschou, S. B. (2025). Assessment of anxiety and behavior disorders among autistic children and youths with intellectual and developmental disabilities. Research in Autism, 121–122, 202550. 10.1016/j.reia.2025.202550

23. Heussler, H., Cohen, J., Buchanan, C., O’Neill, C., Sebree, T., & O’Quinn, S. (2022). An Open-Label, Tolerability and Efficacy Study of ZYN002 (cannabidiol) Administered As A Transdermal Gel To Children And Adolescents With 22Q11.2 Deletion Syndrome (INSPIRE). Journal of Intellectual Disability Research, 66(8–9), 672. Embase. 10.1111/jir.12962

24. Iseger, T. A., & Bossong, M. G. (2015). A systematic review of the antipsychotic properties of cannabidiol in humans. Schizophrenia Research, 162(1–3), 153–161. 10.1016/j.schres.2015.01.033

25. Janssen-Cilag Pty Ltd. (1993). RISPERDAL (risperidone)—Australian Product Information [Australian Product Information]. Janssen–Cilag Pty Ltd.

26. Karhson, D. S., Hardan, A. Y., & Parker, K. J. (2016). Endocannabinoid signaling in social functioning: An RDoC perspective. Translational Psychiatry, 6(9), e905–e905. 10.1038/tp.2016.169

27. Karhson, D. S., Krasinska, K. M., Dallaire, J. A., Libove, R. A., Phillips, J. M., Chien, A. S., Garner, J. P., Hardan, A. Y., & Parker, K. J. (2018). Plasma anandamide concentrations are lower in children with autism spectrum disorder. Molecular Autism, 9, 18.

28. Larsen, C., & Shahinas, J. (2020). Dosage, Efficacy and Safety of Cannabidiol Administration in Adults: A Systematic Review of Human Trials. Journal of Clinical Medicine Research, 12(3), 129–141. 10.14740/jocmr4090

29. Lattanzi, S., Zaccara, G., Russo, E., La Neve, A., Lodi, M. A. M., & Striano, P. (2021). Practical use of pharmaceutically purified oral cannabidiol in Dravet syndrome and Lennox-Gastaut syndrome. Expert Review of Neurotherapeutics, 21(1), 99–110.

30. Manter, M. A., Birtwell, K. B., Bath, J., Friedman, N. D. B., Keary, C. J., Neumeyer, A. M., Palumbo, M. L., Thom, R. P., Stonestreet, E., Brooks, H., Dakin, K., Hooker, J. M., & McDougle, C. J. (2025). Pharmacological treatment in autism: A proposal for guidelines on common co-occurring psychiatric symptoms. BMC Medicine, 23(1), 11. 10.1186/s12916-024-03814-0

31. Norman, G. R., Sloan, J. A., & Wyrwich, K. W. (2003). Interpretation of changes in health-related quality of life: The remarkable universality of half a standard deviation. Medical Care, 41(5), 582–592. 10.1097/01.MLR.0000062554.74615.4C

32. Osborne, A. L., Solowij, N., & Weston-Green, K. (2017). A systematic review of the effect of cannabidiol on cognitive function: Relevance to schizophrenia. Neuroscience & Biobehavioral Reviews, 72, 310–324. 10.1016/j.neubiorev.2016.11.012

33. Papadopoulos, A., Siafaka, V., Tsapara, A., Tafiadis, D., Kotsis, K., Skapinakis, P., & Tzoufi, M. (2023). Measuring parental stress, illness perceptions, coping and quality of life in families of children newly diagnosed with autism spectrum disorder. BJPsych Open, 9(3), e84. 10.1192/bjo.2023.55

34. Parrella, N.-F., Hill, A. T., Enticott, P. G., Barhoun, P., Bower, I. S., & Ford, T. C. (2023). A systematic review of cannabidiol trials in neurodevelopmental disorders. Pharmacology Biochemistry and Behavior, 230, 173607. 10.1016/j.pbb.2023.173607

35. Pretzsch, C. M., Freyberg, J., Voinescu, B., Lythgoe, D., Horder, J., Mendez, M. A., Wichers, R., Ajram, L., Ivin, G., Heasman, M., Edden, R. A. E., Williams, S., Murphy, D. G. M., Daly, E., & McAlonan, G. M. (2019). Effects of cannabidiol on brain excitation and inhibition systems; a randomised placebo-controlled single dose trial during magnetic resonance spectroscopy in adults with and without autism spectrum disorder. Neuropsychopharmacology, 44(8), 1398–1405. 10.1038/s41386-019-0333-8

36. Pretzsch, C. M., Voinescu, B., Mendez, M. A., Wichers, R., Ajram, L., Ivin, G., Heasman, M., Williams, S., Murphy, D. G., Daly, E., & McAlonan, G. M. (2019). The effect of cannabidiol (CBD) on low-frequency activity and functional connectivity in the brain of adults with and without autism spectrum disorder (ASD). Journal of Psychopharmacology, 33(9), 1141–1148. 10.1177/0269881119858306

37. PROMIS Health Organization and Assessment Center. (2014a). PROMIS® Early Childhood Parent-Report v1.0—Anxiety. Northwestern University / NIH.

38. PROMIS Health Organization and Assessment Center. (2014b). PROMIS® Early Childhood Parent-Report v1.0—Sleep Health. Northwestern University / NIH.

39. PROMIS Health Organization and Assessment Center. (2014c). PROMIS® Early Childhood Parent-Report v1.0—Social Relationships. Northwestern University / NIH.

40. Silva Junior, E. A. D., Medeiros, W. M. B., Santos, J. P. M. D., Sousa, J. M. M. D., Costa, F. B. D., Pontes, K. M., Borges, T. C., Neto Segundo, C. E., Andrade E Silva, A. H., Nunes, E. L. G., Alves, N. T., Rosa, M. D. D., & Albuquerque, K. L. G. D. D. (2024). Evaluation of the efficacy and safety of cannabidiol-rich cannabis extract in children with autism spectrum disorder: Randomized, double-blind, and placebo-controlled clinical trial. Trends in Psychiatry and Psychotherapy. 10.47626/2237-6089-2021-0396

41. Silva, L. M. T., & Schalock, M. (2012). Autism Parenting Stress Index: Initial Psychometric Evidence. Journal of Autism and Developmental Disorders, 42(4), 566–574. 10.1007/s10803-011-1274-1

42. Sparrow, S. S., Cicchetti, D. V., & Saulnier, C. A. (2016). Vineland Adaptive Behavior Scales, Third Edition (Vineland-3). Pearson.

43. Sutherland, D. L., Taylor, E. L., Gray, K. M., Hastings, R. P., Allard, A., Carr, J., Griffin, J., McMeekin, N., Randell, E., Russell, D., Willoughby-Richards, B., Wolstencroft, J., & Thompson, P. A. (2025). Estimating a minimum clinically important difference for the Developmental Behaviour Checklist – parent report. Frontiers in Psychiatry, 16, 1612911. 10.3389/fpsyt.2025.1612911

44. Therapeutic Goods Administration (TGA). (2025, April 3). Clarification of requirements to access unapproved medicinal cannabis medicines for paediatric patients. Australian Government Department of Health, Disability and Ageing. https://www.tga.gov.au/resources/publication/publications/clarification-requirements-access-unapproved-medicinal-cannabis-medicines-paediatric-patients

45. Thiele, E. A., Marsh, E. D., French, J. A., Mazurkiewicz-Beldzinska, M., Benbadis, S. R., Joshi, C., Lyons, P. D., Taylor, A., Roberts, C., & Sommerville, K. (2018). Cannabidiol in patients with seizures associated with Lennox-Gastaut syndrome (GWPCARE4): A randomised, double-blind, placebo-controlled phase 3 trial. *Lancet (London*, England), 391(10125), 1085–1096.

46. Thiele-Swift, H. N., & Dorstyn, D.-S. (2024). Anxiety Prevalence in Youth with Autism: A Systematic Review and Meta-analysis of Methodological and Sample Moderators. Review Journal of Autism and Developmental Disorders. 10.1007/s40489-023-00427-w

47. VanDolah, H. J., Bauer, B. A., & Mauck, K. F. (2019). Clinicians’ Guide to Cannabidiol and Hemp Oils. Mayo Clinic Proceedings, 94(9), 1840–1851. 10.1016/j.mayocp.2019.01.003

48. Varni, J. W., Seid, M., & Rode, C. A. (1999). The PedsQL: Measurement model for the Pediatric Quality of Life Inventory. Medical Care, 37(2), 126–139. 10.1097/00005650-199902000-00003

49. Veenstra-VanderWeele, J., Cook, E. H., King, B. H., Zarevics, P., Cherubini, M., Walton-Bowen, K., Bear, M. F., Wang, P. P., & Carpenter, R. L. (2017). Arbaclofen in Children and Adolescents with Autism Spectrum Disorder: A Randomized, Controlled, Phase 2 Trial. Neuropsychopharmacology, 42(7), 1390–1398. 10.1038/npp.2016.237

50. Wei, D., Dinh, D., Lee, D., Li, D., Anguren, A., Moreno-Sanz, G., Gall, C. M., & Piomelli, D. (2016). Enhancement of Anandamide-Mediated Endocannabinoid Signaling Corrects Autism-Related Social Impairment. Cannabis and Cannabinoid Research, 1(1), 81–89. 10.1089/can.2015.0008

51. Zamberletti, E., Gabaglio, M., & Parolaro, D. (2017). The Endocannabinoid System and Autism Spectrum Disorders: Insights from Animal Models. International Journal of Molecular Sciences, 18(9), 1916. 10.3390/ijms18091916

