## Supplemental Materials for "Effects of Cannabidiol on Social Relating, Anxiety, and Parental Stress in Autistic Children: A Randomized Controlled Crossover Trial"

**Supplementary Methods S1. Parent Instructions for Administration**

Parents/guardians were provided in-person training on the administration of the investigational product. The researcher demonstrated the following procedure during the first visit:

1. Dose calculation
   - The daily dose was calculated individually for each child based on the formula: Dose (mg/day) = 10 mg x [child's weight in kg]
   - For example, a 30 kg child received 300 mg/day.
2. Demonstration
   - Parents were shown the product bottle and how to draw up the correct dose using the provided measuring syringe.
   - The researcher demonstrated measuring the dose accurately to the nearest 0.1 mL.
3. Timing of doses
   - Parents were advised to divide the total daily dose into two administrations:
   - First dose: typically before school or in the morning.
   - Second dose: after school/in the evening, before bedtime.
   - Parents were encouraged to choose times that best suited the child's daily routine, while maintaining consistency each day.
4. Additional guidance
   - Shake the bottle well before each dose.
   - Administer the dose directly or mix with a small amount of soft food/drink if needed.
   - Record administration times in the daily treatment log provided via web link.
   - Contact the research team with any concerns or if a dose was missed.

**Supplementary Table S1. Baseline equivalence by sequence (A: CBD-Placebo; B: Placebo-CBD)**

| Outcome | Sequence A(Period 1 = CBD - Placebo)Mean (SD) | Sequence B(Period 1 = Placebo - CBD)Mean (SD) | *p* |
|---|---|---|---|
| **Age (years)** | 9.5 (2.0) | 9.7 (2.1) | .73 |
| **Sex (male / female)** | 10/7 | 8/4 | .88 |
| **SRS-2** | | | |
| Social Awareness | 15.1 (4.85) | 14.9 (4.69) | .26 |
| Social Cognition | 22.6 (7.41) | 22.2 (7.48) | .44 |
| Social Communication | 39.5 (12.7) | 37.6 (13.5) | .15 |
| Social Motivation | 17.6 (6.92) | 17.7 (5.87) | .41 |
| Restricted/Repetitive | 25.6 (7.00) | 24.8 (7.90) | .32 |
| **Total (Composite)** | 94.9 (29.6) | 92.4 (29.9) | .23 |
| **RBS-R** | | | |
| Stereotypic | 8.36 (6.03) | 8.10 (5.65) | .53 |
| Injurious | 4.29 (3.71) | 3.77 (3.61) | .25 |
| Compulsive | 4.11 (4.38) | 3.83 (3.78) | .61 |
| Ritualistic | 11.30 (7.72) | 10.30 (7.66) | .46 |
| Restricted Interests | 3.93 (2.64) | 3.80 (2.41) | .53 |
| **Total** | 31.9 (20.9) | 29.8 (18.8) | .37 |
| **PedsQL** | | | |
| Physical | 15.3 (6.66) | 13.3 (6.49) | .62 |
| Emotional | 11.6 (3.98) | 11.4 (4.25) | .62 |
| Social | 12.1 (4.56) | 11.5 (5.00) | .43 |
| School | 11.4 (4.06) | 10.8 (4.17) | .24 |
| **Total** | 50.3 (14.8) | 47.0 (16.6) | .37 |
| **APSI Total** | 23.9 (11.5) | 25.1 (12.3) | .84 |
| **PROMIS (raw)** | | | |
| Anxiety | 20.7 (7.51) | 21.2 (7.50) | .65 |
| Sleep Health | 21.4 (9.14) | 18.7 (7.79) | .72 |
| Social Relationships | 20.0 (5.21) | 20.2 (5.03) | .32 |
| **BRIEF-2** | | | |
| Inhibit | 19.5 (3.89) | 18.6 (5.07) | .72 |
| Self-Monitor | 10.3 (1.96) | 9.73 (2.49) | .20 |
| Shift | 20.7 (3.35) | 19.8 (4.81) | .52 |
| Emotional Control | 22.9 (3.35) | 21.3 (5.87) | .22 |
| Initiate | 12.9 (2.23) | 12.4 (3.32) | .67 |
| Working Memory | 20.6 (3.39) | 19.7 (5.13) | .82 |
| Planning | 20.6 (2.87) | 19.4 (5.11) | .42 |
| Task Monitor | 12.7 (2.16) | 12.4 (3.06) | .75 |

| Outcome | Sequence A(Period 1 = CBD - Placebo)Mean (SD) | Sequence B(Period 1 = Placebo - CBD)Mean (SD) | *p* |
|---|---|---|---|
| **Age (years)** | 9.5 (2.0) | 9.7 (2.1) | .73 |
| **Sex (male / female)** | 10/7 | 8/4 | .88 |
| Organization | 14.5 (3.08) | 13.9 (4.15) | .85 |
| **Vineland-3** | | | |
| Adaptive Behavior Composite | 68.0 (15.9) | 69.0 (14.0) | .56 |
| Communication | 68.1 (20.6) | 69.4 (18.5) | .60 |
| Daily-Living Skills | 70.0 (17.7) | 70.3 (16.9) | .80 |
| Socialization | 64.8 (16.7) | 67.4 (15.4) | .30 |
| Motor Skills† | 70.8 (23.0) | 74.5 (21.3) | .31 |
| **DBC-2 (T-scores)** | | | |
| Total | 64.4 (12.8) | 66.2 (13.0) | .44 |
| Disruptive | 59.4 (11.9) | 61.6 (11.4) | .71 |
| Self-Absorbed | 60.3 (12.8) | 61.6 (13.0) | .69 |
| Communication | 66.2 (6.19) | 65.6 (5.63) | .35 |
| Anxiety | 64.9 (11.3) | 66.3 (11.5) | .21 |
| Social Relating | 60.4 (12.4) | 61.9 (10.9) | .59 |

**Supplementary Figure S1.** Model-estimated means (95% confidence interval [CI]) by treatment and period for the primary outcome (Social Responsiveness Scale [SRS-2] Total) and SRS-2 subscales (SCI, Communication, Cognition, Motivation, Awareness) to show baseline comparability (T0 vs T2) and 12-week endpoints (T1 vs T3). Active/CBD is shown in light blue and Placebo in dark blue in each period.

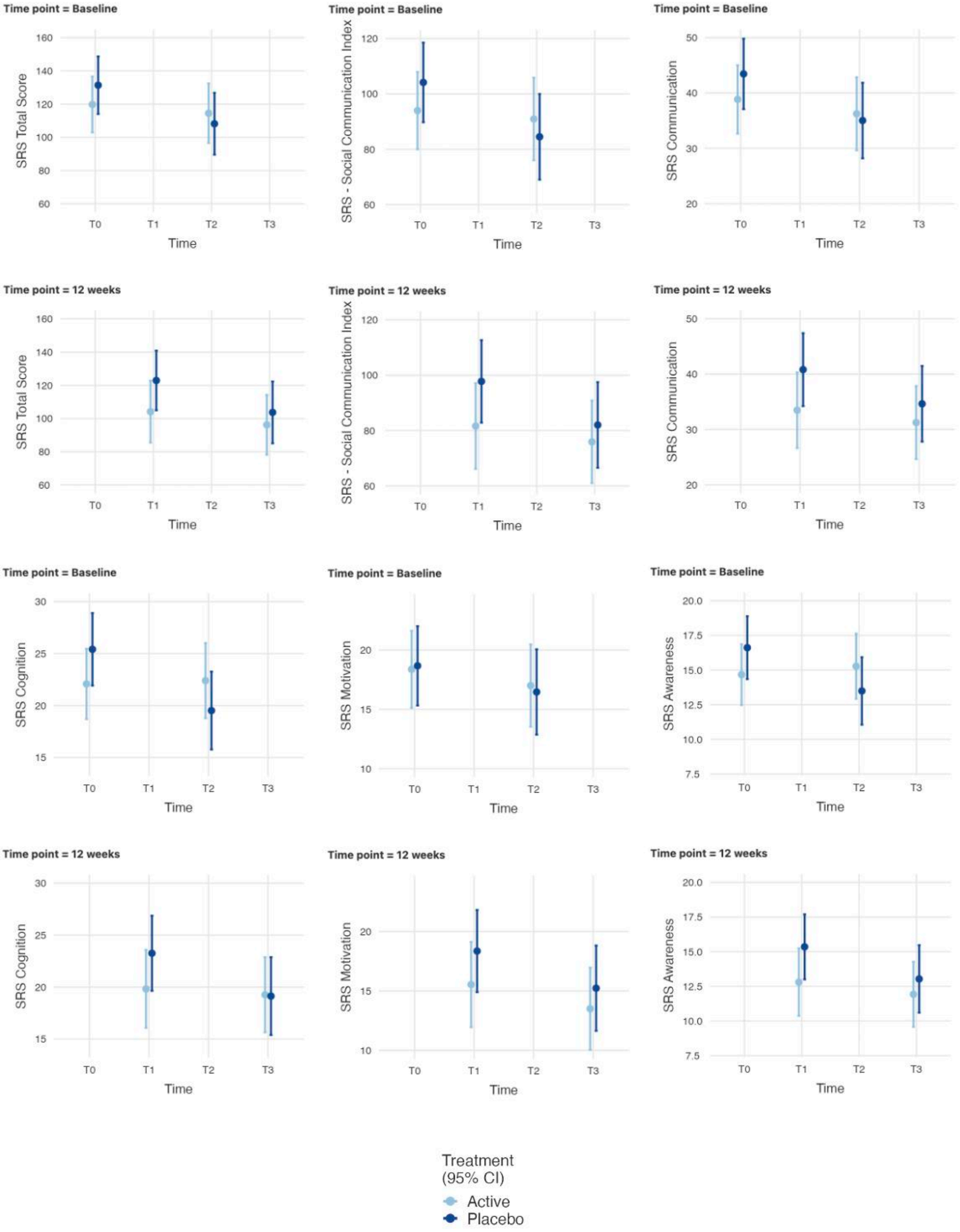

**Supplementary Figure S2.** Model-estimated means (95% confidence interval [CI]) by treatment and period for secondary outcomes (DBC-2 Social Relating/Anxiety; Vineland-3 ABC, Communication, Daily Living, Socialization; Autism Parenting Stress Index [APSI]; PROMIS Anxiety/Social) to show baseline comparability (T0 vs T2) and 12-week endpoints (T1 vs T3). Active/CBD is shown in light blue and Placebo in dark blue in each period.

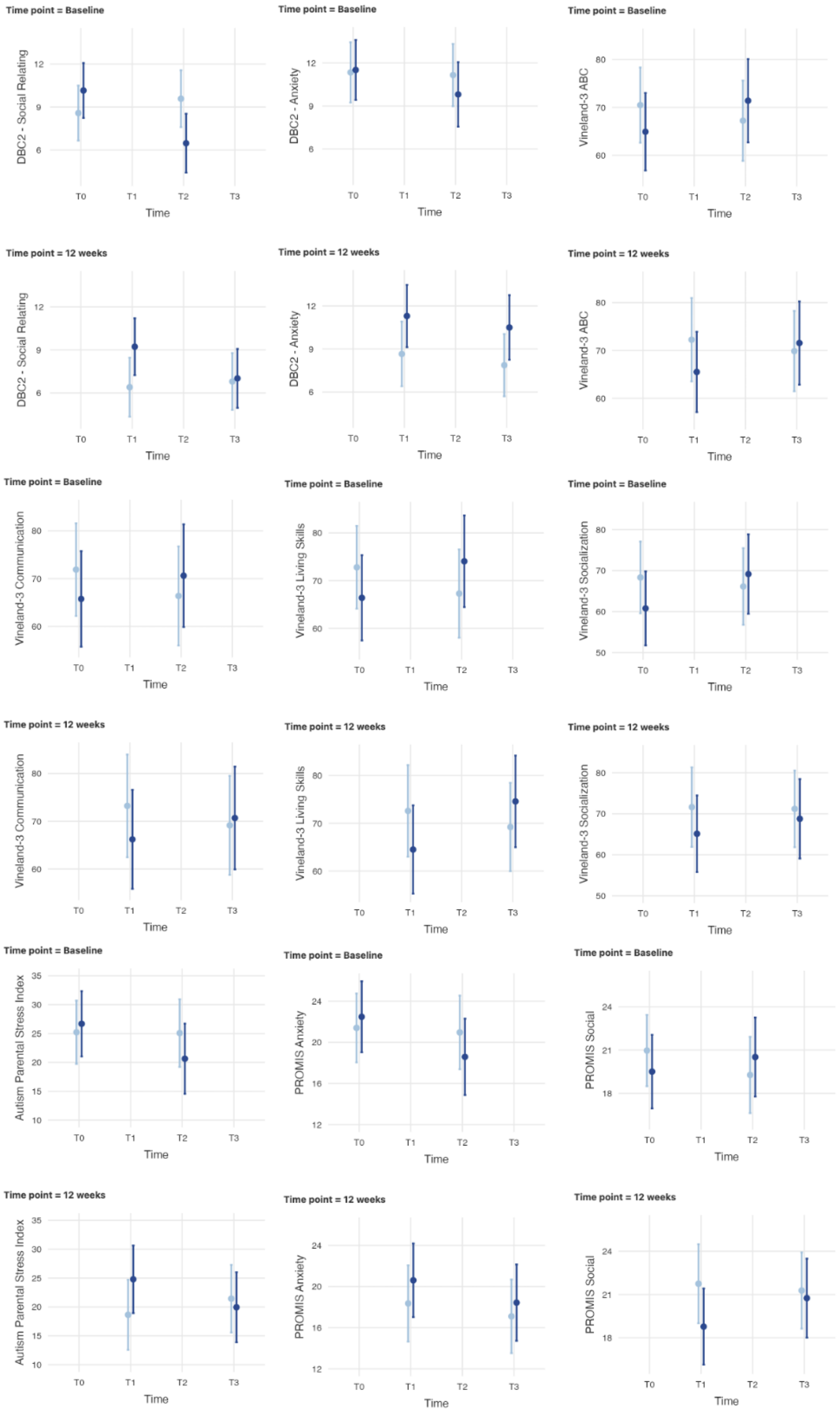

**Supplemental Table S2. Exploratory analyses on remaining behavioral outcome measures from baseline to 12-weeks.**

| Measure | Intervention | Time Point | Mean (SD) | F(df) | β | SE | ηp² | *p*-value |
|---|---|---|---|---|---|---|---|---|
| BRIEF - Emotional Control | Placebo | Baseline | 22.8 (3.36) | 0.008 (1, 78.9) | 0.0111 | 1.321 | 0.000 | .993 |
| | | 12 weeks | 20.7 (5.78) | | | | | |
| | CBD | Baseline | 22.0 (4.41) | | | | | |
| | | 12 weeks | 19.9 (4.44) | | | | | |
| BRIEF - Inhibition | Placebo | Baseline | 19.3 (3.90) | 0.210 (1, 78.9) | 0.2605 | 1.24 | 0.003 | .834 |
| | | 12 weeks | 17.8 (4.98) | | | | | |
| | CBD | Baseline | 19.3 (3.78) | | | | | |
| | | 12 weeks | 18.0 (3.56) | | | | | |
| BRIEF - Self-Monitoring | Placebo | Baseline | 10.3 (2.00) | 0.539 (1, 78.7) | -0.358 | 0.665 | 0.007 | .592 |
| | | 12 weeks | 9.93 (2.66) | | | | | |
| | CBD | Baseline | 10.1 (1.74) | | | | | |
| | | 12 weeks | 9.37 (2.19) | | | | | |
| BRIEF - Shifting | Placebo | Baseline | 20.7 (3.41) | 0.539 (1, 78.7) | -0.358 | 0.665 | 0.007 | .592 |
| | | 12 weeks | 18.8 (5.13) | | | | | |
| | CBD | Baseline | 20.5 (3.13) | | | | | |
| | | 12 weeks | 18.6 (3.36) | | | | | |
| Developmental Behavior Checklist (DBC-2) Total | Placebo | Baseline | 76.1 (31.7) | 3.15 (1, 78.4) | -7.62 | 2.42 | 0.038 | **.002*** |
| | | 12 weeks | 74.8 (28.5) | | | | | |
| | CBD | Baseline | 80.2 (30.1) | | | | | |
| | | 12 weeks | 61.4 (21.3) | | | | | |
| Pediatric Quality of Life Inventory (PedQL) Total | Placebo | Baseline | 50.5 (15.1) | 0.348 (1, 78.7) | 1.09 | 3.12 | 0.004 | .729 |
| | | 12 weeks | 46.4 (15.3) | | | | | |

| | | | | | | | | |
|---|---|---|---|---|---|---|---|---|
| | CBD | Baseline | 47.5 (17.1) | | | | | |
| | | 12 weeks | 44.6 (11.3) | | | | | |
| PROMIS Sleep | Placebo | Baseline | 21.3 (9.30) | 1.325 (1, 78.9) | -2.704 | 2.04 | 0.017 | .189 |
| | | 12 weeks | 19.7 (9.20) | | | | | |
| | CBD | Baseline | 23.0 (8.25) | | | | | |
| | | 12 weeks | 18.7 (7.79) | | | | | |
| Repetitive Behavior Scale (RBS) Total | Placebo | Baseline | 32.7 (20.9) | 0.707 (1, 78.2) | -2.15 | 3.91 | 0.008 | .585 |
| | | 12 weeks | 30.0 (17.4) | | | | | |
| | CBD | Baseline | 30.2 (19.4) | | | | | |
| | | 12 weeks | 25.9 (13.0) | | | | | |

**Supplementary Table S3. CONSORT Harms 2022 integrated into CONSORT 2010 items checklist of information to include when reporting a randomised trial**

| Section/Topic | Item No | Checklist item | Reported on page No |
|---|---|---|---|
| **Title and abstract** | | | |
| | 1a | Identification as a randomised trial in the title | 83 |
| | 1b | Structured summary of trial design, methods, results of outcomes of benefits and harms, and conclusions (for specific guidance see CONSORT for abstracts) | 83 |
| **Introduction** | | | |
| Background and objectives | 2a | Scientific background and explanation of rationale | 84 |
| | 2b | Specific objectives or hypotheses for outcomes benefits and harms | 84 |
| **Methods** | | | |
| Trial design | 3a | Description of trial design (such as parallel, factorial) including allocation ratio | 88 |
| | 3b | Important changes to methods after trial commencement (such as eligibility criteria), with reasons | N/A |
| Participants | 4a | Eligibility criteria for participants | 89 |
| | 4b | Settings and locations where the data were collected | 88 |
| Interventions | 5 | The interventions for each group with sufficient details to allow replication, including how and when they were actually administered | 92 |
| Outcomes | 6a | Completely defined pre-specified primary and secondary outcome measures for both benefits and harms, including how and when they were assessed | 94 |
| | 6b | Any changes to trial outcomes after the trial commenced, with reasons | N/A |
| | 6c | Describe if and how non-prespecified outcomes of benefits and harms were identified, including any selection criteria, if applicable | N/A |
| Sample size | 7a | How sample size was determined | 95 |
| | 7b | When applicable, explanation of any interim analyses and stopping guidelines | N/A |
| Randomisation: | | | |
| Sequence generation | 8a | Method used to generate the random allocation sequence | 93 |
| | 8b | Type of randomisation; details of any restriction (such as blocking and block size) | 93 |
| Allocation concealment mechanism | 9 | Mechanism used to implement the random allocation sequence (such as sequentially numbered containers), describing any steps taken to conceal the sequence until interventions were assigned | 93 |
| Implementatio n | 10 | Who generated the random allocation sequence, who enrolled participants, and who assigned participants to interventions | 93 |
| Blinding | 11a | If done, who was blinded after assignment to interventions (e.g., participants, care providers, those assessing outcomes of benefits and harms) and how | 93 |
| | 11b | If relevant, description of the similarity of interventions | 92 |
| Statistical methods | 12a | Statistical methods used to compare groups for primary and secondary outcomes of both benefits and harms | 97 |
| | 12b | Methods for additional analyses, such as subgroup analyses and adjusted analyses | 97, 98 |
| **Results** | | | |
| Participant flow (a diagram is | 13a | For each group, the numbers of participants who were randomly assigned, received intended treatment, and were analysed for outcomes of benefits and harms | 99 |

| Section/Topic | Item No | Checklist item | Reported on page No |
|---|---|---|---|
| strongly recommended) | 13b | For each group, losses and exclusions after randomisation, together with reasons | 99 |
| Recruitment | 14a | Dates defining the periods of recruitment and follow-up for outcomes of benefits and harms | N/A |
| | 14b | Why the trial ended or was stopped | N/A |
| Baseline data | 15 | A table showing baseline demographic and clinical characteristics for each group | 90 |
| Numbers analysed | 16 | For each group, number of participants (denominator) included in each analysis and whether the analysis was by original assigned groups and if any exclusions were made | 105 |
| Outcomes and estimation | 17a | For each primary and secondary outcome of benefits and harms, results for each group, and the estimated effect size and its precision (such as 95% confidence interval) | 105-108 |
| | 17a2 | For outcomes omitted from the trial report (benefits and harms), provide rationale for not reporting and indicate where the data on omitted outcomes can be accessed | 99 |
| | 17b | Presentation of both absolute and relative effect sizes is recommended, for outcomes of benefits and harms | 105-108 |
| | 17c | Report zero events if no harms were observed | N/A |
| Ancillary analyses | 18 | Results of any other analyses performed, including subgroup analyses and adjusted analyses, distinguishing pre-specified from exploratory | 196-198 |
| Harms | 19 | All important harms or unintended effects in each group (for specific guidance see CONSORT for harms) | 104 |
| **Discussion** | | | |
| Limitations | 20 | Trial limitations, addressing sources of potential bias related to the approach to collecting or reporting data on harms, imprecision, and, if relevant, multiplicity or selection of analyses | 111-112 |
| Generalisability | 21 | Generalisability (external validity, applicability) of the trial findings | 112 |
| Interpretation | 22 | Interpretation consistent with results, balancing benefits and harms, and considering other relevant evidence | 109-112 |
| **Other information** | | | |
| Registration | 23 | Registration number and name of trial registry | 88 |
| Protocol | 24 | Where the full trial protocol and other relevant documents can be accessed, including additional data on harms | 88 |
| Funding | 25 | Sources of funding and other support (such as supply of drugs), role of funders | 114 |

Note: Adapted from Schulz (2010) to integrate items of CONSORT Harms 2022 (Junqueira 2022) [https://creativecommons.org/licenses/by/2.0/]. CONSORT items 1b, 2b, 6a, 11a, 12a, 13a, 14a, 16a, 17a, 17b, 18, 20 and 24 of were modified to incorporate elements relevant to the reporting of harms. Two new items were added (item 6c and 17a2). Please see the CONSORT Harms 2022 statement for additional details (Junqueira 2022).

**References**

Junqueira DR, Zorzela L, Golder S, Loke Y, Gagnier JJ, Julious SA, Li T, Mayo-Wilson E, Pham B, Phillips R, Santaguida P, Scherer RW, Gøtzsche PC, Moher D, Ioannidis JPA and Vohra S on behalf of the CONSORT Harms Group. CONSORT Harms 2022 statement, explanation, and elaboration: updated guideline for the reporting of harms in randomised trials. *BMJ* 2023 **381:** e073725 DOI 10.1136/bmj-2022-073725

Schulz KF, Altman DG, Moher D, for the CONSORT Group. CONSORT 2010 Statement: updated guidelines for reporting parallel group randomised trials. *BMJ* 2010 **340:**c332 doi: 10.1136/bmj.c332.

### Supplementary Form S1. Daily Treatment Log

#### CBD Autism: Treatment Log

Start of Block: Default Question Block

Q1 Participant Identification Number

________________________________________________________________

Q2 Date

________________________________________________________________

Q3 Did your child take the oil today?

- ○ Yes
- ○ Partially. Please comment how much was taken: ________________________________________________
- ○ No

Q4 How many times was the oil taken?

- ○ Once
- ○ Twice
- ○ Three times
- ○ More than three times
- ○ Not taken today.

Q5 What time/s of the day did they take the oil?

- ☐ 6am - 10am
- ☐ 10am - 12pm
- ☐ 12pm - 2pm
- ☐ 2pm - 4pm
- ☐ 4pm - 6pm
- ☐ 6pm - 8pm
- ☐ 8pm - 10pm
- ☐ 10pm - 12am
- ☐ 12am - 6am
- ☐ None taken today

Q6 <h4>How many times was the oil taken with food?</h4>

- ○ Once
- ○ Twice
- ○ Three times
- ○ More than three times
- ○ Not taken with food today.

Q7 If the oil was taken with food, please write what food it was consumed with:

________________________________________________________________

Q8 Any comments about administration?

________________________________________________________________

Q9 How would you describe your child's appetite today?

| | No appetite | Mild appetite | Healthy appetite | Big appetite | Extremely big appetite |
|---|---|---|---|---|---|
| Desire to eat | ○ | ○ | ○ | ○ | ○ |

Q10 The 'Feelings Thermometer' can be used to help children identify their feelings. See below for descriptions for 'Zones':

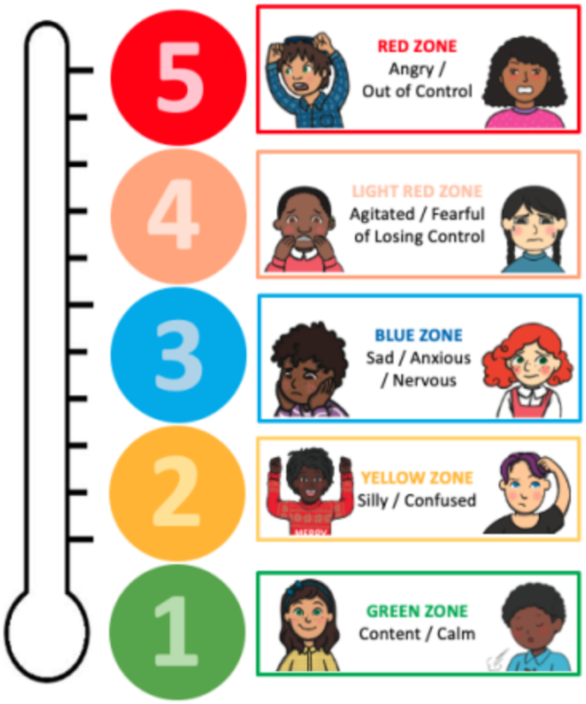


Q11 Which of the 'Zones' has your child experienced today?

- ☐ Green (e.g. positive/calm)
- ☐ Yellow (e.g. excited/silly/confused)
- ☐ Blue (e.g. unhappy/withdrawn/nervous)
- ☐ Light red (e.g. agitated/frustrated)
- ☐ Red (e.g. mad/yelling)

Q12 Any issues to report?

- ○ Yes ________________________________________________
- ○ No
- ○ 6

Q13 Any further comments?

________________________________________________

End of Block: Default Question Block
